# Selective degradation of pathogenic autoantibodies by lysosomal targeting chimeras for the treatment of myasthenia gravis

**DOI:** 10.1101/2025.11.18.25340499

**Authors:** Nicholas A. Lind, Christina L. Collins, Craig Park, Rozhan Khaleghi, Sarah M. Totten, Diana Li, Tao Chen, Rachel M. Lieser, Thomas-Toan Tran, Carly Cheung, Gianvito Masi, Richard J. Nowak, Steven T. Staben, Eric D. Turtle, Darrin A. Lindhout, Kevin C. O’Connor, Sarah M. McWhirter, Jeffrey S. Iwig

## Abstract

Myasthenia gravis, an autoimmune disease characterized by muscle weakness that can manifest in bulbar dysfunction and respiratory distress, is driven in a subset of patients by autoantibodies against muscle-specific receptor tyrosine kinase (MuSK MG). MuSK MG and other autoantibody mediated disorders are generally treated with broad immunosuppressive approaches such as corticosteroids, plasmapheresis, B cell depletion, or FcRn inhibition, which can impact patient care via systemic side effects, variable responsiveness, and increased susceptibility to infection. We developed a lysosomal targeting chimera (LYTAC) therapeutic (MuSK LYTAC) that combines a MuSK antigen “Bait” capable of selectively binding to the disease-driving antibodies with asialoglycoprotein receptor (ASGPR) ligands that target the resulting immune complexes for degradation by ASGPR-expressing hepatocytes. Our optimized MuSK Bait effectively depleted pathogenic antibodies from the serum of 10 donors with MuSK MG, supporting broad applicability across the patient population. In vitro assays using both Hep G2 cells and primary human hepatocytes demonstrated that MuSK LYTAC is pharmacologically active, promoting uptake and lysosomal degradation of autoantibodies through an ASGPR-dependent mechanism. Mouse pharmacodynamic and passive immunization disease models showed that MuSK LYTAC rapidly eliminates autoantibodies from the serum and alleviates related disease symptoms, without impacting circulating IgG levels. These results support the promise of Autoantibody Bait LYTACs (ABLates) as precision treatments for MuSK MG and other autoantibody-driven diseases that avoid generalized immunosuppression.

## Introduction

Autoimmune diseases arise from abnormal immune responses against self-antigens, that lead to tissue damage and dysfunction. While certain autoimmune diseases involve broad autoantibody responses against multiple distinct antigens, myasthenia gravis (MG) is a paradigmatic example of a B-cell driven disorder mediated by autoantibodies targeting a single dominant autoantigen (1). Autoantibodies can play a central role in disease pathogenesis through diverse mechanisms such as disrupting cellular signaling pathways, inducing cell lysis and promoting inflammation (2–4). Patients with MG suffer from muscle weakness and fatigue due to autoantibodies that target key transmembrane proteins at the neuromuscular junction (NMJ) (5). While most MG patients test positive for autoantibodies targeting the nicotinic acetylcholine receptor (AChR), 5-10% harbor autoantibodies to the muscle-specific receptor tyrosine kinase (MuSK), a protein that orchestrates NMJ development and maintenance (1). MuSK MG requires specialized neuromuscular/ neurologic care due to the frequency of bulbar and respiratory muscle involvement leading to severe and life-threatening swallowing and breathing difficulty in some cases, highlighting the complexity of managing this subtype of MG (6). Multiple lines of evidence support the pathogenic capacity of anti-MuSK autoantibodies (7–9). As these autoantibodies are predominantly of the IgG4 subclass, they engage in a stochastic process known as “fragment antigen binding (Fab)-arm exchange” (10, 11) in which MuSK IgG4 autoantibodies randomly exchange half-molecules (one heavy and one light chain) with other IgG4 antibodies, thus retaining a single MuSK binding arm. Studies have shown that the resulting “functional monovalency”, a unique property of IgG4, augments the pathogenic effects of MuSK autoantibodies (9, 12, 13). Functionally monovalent MuSK IgG4 autoantibodies block the binding between MuSK and the agrin/low-density lipoprotein receptor-related protein 4 complex, thereby inhibiting a signaling cascade required to maintain neuromuscular synapses.

Many MuSK MG patients are treated with high doses of corticosteroids and non-steroidal immunosuppressants yet remain symptomatic after first-line therapy and require therapeutic anti-CD20 B cell depletion with rituximab to achieve clinical stability (14, 15). Although effective, disease relapses can occur during B-cell reconstitution (12). Symptom re-exacerbation during relapses may be severe, necessitating hospitalization and further intensive management for long-term disease control (6). These limitations highlight the need for more selective therapies to control disease with fewer systemic side effects.

Precision immunology approaches, where pathogenic B cells or autoantibodies are selectively targeted, hold great promise for controlling MuSK MG without widespread suppression of immune function. Initial attempts to treat autoantibody-mediated disorders with targeted immunotherapies, however, have not been successfully adapted to the clinic. For example, antigen-specific T cell approaches using chimeric autoantibody receptor T cell (CAAR-T) therapy have shown promising preclinical results in treating MuSK MG and pemphigus vulgaris (16). However, in Phase 1 trials, CAAR-T treatment led to limited autoantibody reduction and serious adverse events in some cases (17).

Extracellular targeted protein degradation (eTPD) therapies offer a way to specifically eliminate disease-driving MuSK autoantibodies from circulation (18, 19). eTPD approaches, such as lysosomal targeting chimeras (LYTACs) utilize heterobifunctional molecules that induce proximity between an extracellular target protein and a cell-surface internalizing receptor to promote target trafficking to the lysosome for degradation (18). LYTACs and similar approaches have been developed that utilize the asialoglycoprotein receptor (ASGPR) expressed on hepatocytes as an internalizing receptor to drive rapid and deep removal of target proteins from circulation (20–22). ASGPR is a high capacity, rapidly internalizing, liver-specific protein that naturally removes N-acetylgalactosamine or galactose bearing proteins from the serum and delivers them for lysosomal degradation (21, 23–26). LYTACs can be designed to degrade a polyclonal population of autoantibodies to a single antigen by utilizing an antigen “bait” as the target binder tethered to a ligand directed to ASGPR. These Autoantibody Bait LYTACs (ABLates) are autoantibody-specific and could be capable of removing all pathogenic autoantibodies across Ig isotypes (e.g., IgG, IgA, IgM) and subclasses (IgG1-IgG4; IgA1-IgA2).

Here we describe a novel ABLate therapeutic, MuSK LYTAC, designed to selectively remove pathogenic MuSK-specific autoantibodies for the treatment of MuSK MG. The MuSK LYTAC antigen bait contains the first two domains of the extracellular region of MuSK, which include the most relevant autoantigenic epitopes (7, 27). We demonstrate the broad applicability of MuSK LYTAC by its ability to target clinically relevant MuSK autoantibodies from a diverse panel of patient sera. We find that MuSK LYTAC internalizes and degrades human MuSK MG autoantibodies in vitro, rapidly removes circulating human MuSK MG auto-antibodies in mice, and protects against disease in a mouse model of MG. The findings presented here highlight MuSK LYTAC as a promising new treatment for MuSK MG designed to eliminate the specific pathogenic autoantibodies responsible for MuSK MG without causing global immune suppression. Our data suggest that the ABLate treatment approach has the potential to be broadly applicable to other antigen-specific autoantibody-driven diseases.

## Results

### MuSK LYTAC design and binding properties

We designed an ABLate containing a portion of the MuSK extracellular region, capable of high affinity autoantibody binding, conjugated to internalizing receptor ligands, which enable the delivery of autoantibodies to the lysosome for degradation. The MuSK ectodomain is comprised of three immunoglobulin-like (Ig) domains and a C-terminal Friz-zled-like domain (Fzd) (Fig. 1A). Previous studies indicated that most patient anti-MuSK antibodies bind to the Ig1 and Ig2 extracellular domains of MuSK (7, 28, 29). We therefore generated a monovalent fusion protein containing the Ig1 and Ig2 domains to capture pathogenic MuSK antibodies, linked via a flexible (Gly4-Ser)3 linker to human serum albumin (HSA) (referred to hereafter as MuSK Bait). HSA provides a purification handle and additional conjugation sites for internalizing receptor ligands (Fig. 1A). The purified fusion protein demonstrated a well-resolved peak by analytical size exclusion chromatography, with a retention volume consistent with a monomeric species (Fig. 1B). To generate MuSK LYTAC, lysine residues of MuSK Bait were stochastically conjugated to a chemical linker displaying high-affinity, trivalent ASGPR ligands (Fig. 1A, Fig. 1C). The average conjugation ratio for MuSK LYTAC was determined to be 5.67 and 5.62 linker-ligands/ protein, as measured by hydrophilic interaction chromatography (HILIC) and mass spectrometry, respectively (fig. S1). Conjugation site mapping of MuSK LYTAC indicated that lysine residues on both MuSK and HSA domains were labeled. Importantly, the conjugation site distribution was proportional to the number of lysines on each protein component, with the highest abundance conjugated peptides originating from HSA. This analysis indicated that there was no site enrichment on MuSK lysines (table S1).

**Figure 1.**
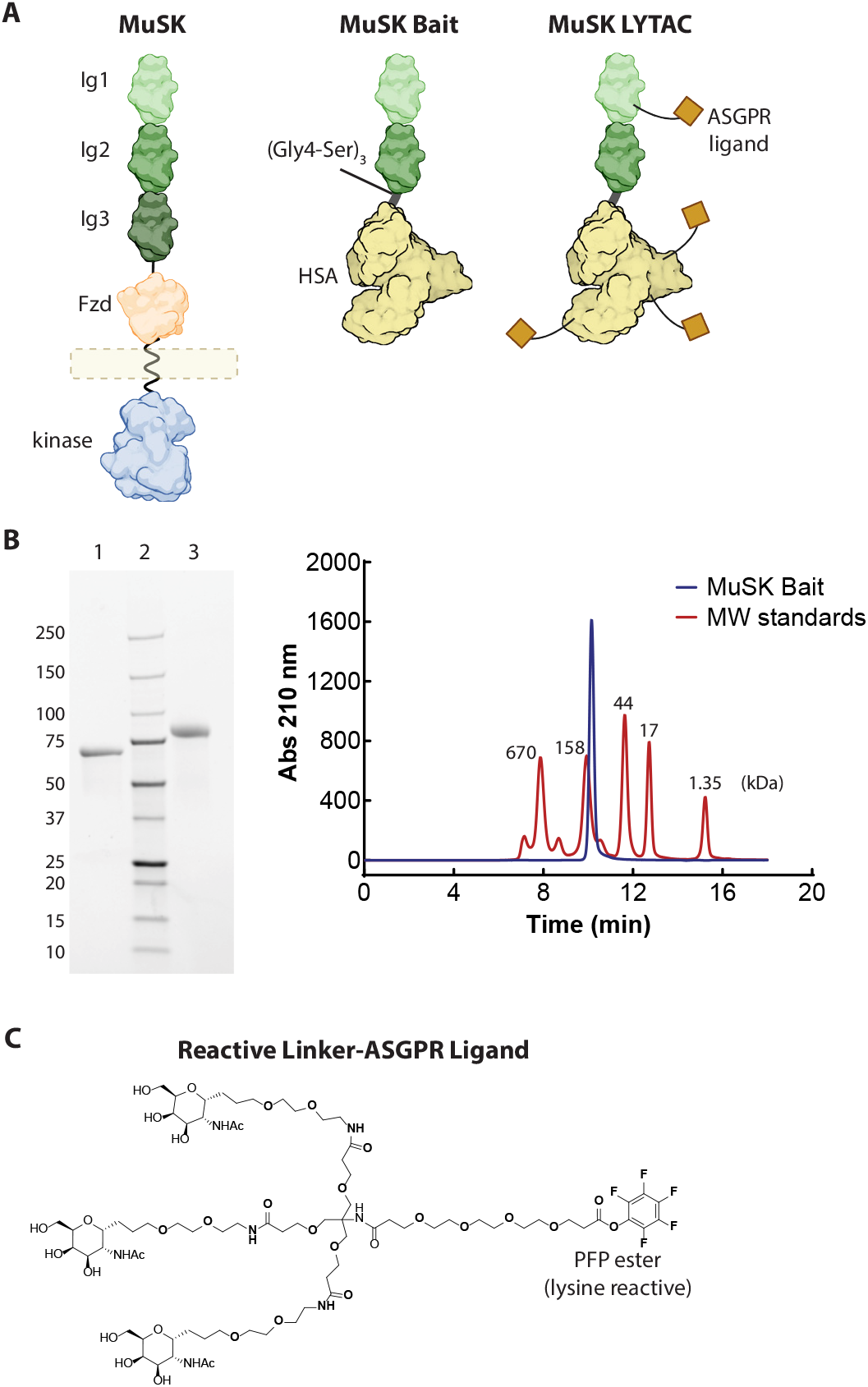
Design of MuSK LYTAC. **(A)** MuSK Bait contains the Ig1 and Ig2 domains of MuSK fused to HSA through a (Gly4-Ser)3 linker. MuSK LYTAC is MuSK Bait conjugated to a linker-ASGPR ligand. **(B)** Nonreducing (lane 1) and reducing (lane 3) SDS-polyacrylamide gel of MuSK Bait after two-step purification with molecular weight marker (lane 2) and indicated sizes (left). Analytical size exclusion chromatography trace of purified MuSK Bait shown relative to MW standards (kDa) (right). **(C)** Reactive linker-ASGPR ligand, activated as a pentafluorophenyl (PFP) ester, used to conjugate MuSK Bait to generate MuSK LYTAC.

We tested the ability of MuSK LYTAC and MuSK Bait to bind previously described monoclonal autoantibodies derived from MuSK MG patients (referred to throughout as MuSK antibodies) (7, 9) by surface plasmon resonance (SPR) and a competition-based time-resolved fluorescence resonance energy transfer (TR-FRET) assay. Antibody binders to both the Ig1 (13-3B5, 11-3F6) and Ig2 (MuSK1A, MuSK1B) domains of MuSK were produced recombinantly. Binding of bivalent or monovalent antibody formats was evaluated, given the known propensity of pathogenic MuSK antibodies to be of the IgG4 subclass (10, 13, 30, 31). IgG4 antibodies are monovalent and bispecific due to Fab-arm exchange (11, 32–34). Monovalent MuSK antibodies were generated by incorporation of an anti-keyhole limpet hemocyanin (KLH) antibody arm (13-3B5-KLH, 11-3F6-KLH and MuSK1A-KLH). MuSK1B-KLH could not be assembled with sufficient purity and was therefore not included in the studies described below. Both MuSK LYTAC and MuSK Bait bound bivalent and monovalent MuSK antibodies targeting Ig1 or Ig2 domains with pM affinities and with good correlation across the two different methods, except for MuSK1B, which showed eight-fold weaker binding by SPR (Table 1, fig. S2 and S3). The IC50 values for the monovalent antibodies in the TR-FRET assay were two-to four-fold higher than the corresponding bivalent antibodies, while the affinity differences between MuSK LYTAC and MuSK Bait for the same antibody were generally less than two-fold. These results demonstrate that ASGPR-ligand conjugation of MuSK Bait does not significantly impact antibody binding and supports the use of MuSK LYTAC as a high-affinity MuSK antibody binder.

**Table 1.** MuSK Bait and MuSK LYTAC binding to MuSK antibodies.

| Antibody | Interaction Site | Valency | MuSK Bait SPR (pM) | MuSK LYTAC SPR (pM) | MuSK Bait TR-FRET (pM) | MuSK LYTAC TR-FRET (pM) |
| --- | --- | --- | --- | --- | --- | --- |
| 13-3B5 | Ig1 | bivalent | 28±4 | 49±17 | 23±8 | 36±7 |
| 13-3B5-KLH | Ig1 | monovalent | nt | nt | 77±8 | 123±11 |
| 11-3F6 | Ig1 | bivalent | 28±6 | nt | 24±3 | 54±11 |
| 11-3F6-KLH | Ig1 | monovalent | nt | nt | 77±8 | 152±32 |
| MuSK1A | Ig2 | bivalent | 27±10 | nt | 17±3 | 18±1 |
| MuSK1A-KLH | Ig2 | monovalent | nt | nt | 44±24 | 37±12 |
| MuSK1B | Ig2 | bivalent | 203±81 | nt | 24±9 | 17±3 |

To degrade MuSK antibodies, MuSK LYTAC must simultaneously bind the antibody and internalizing receptor. The ability of MuSK LYTAC to form ternary complexes with antibody and ASGPR was tested using a TR-FRET assay with Alex-Fluor 647 (AF647)-labeled antibody and a biotin-labeled soluble ASGPR fragment. Titration of MuSK LYTAC produced the expected bell-shaped dose-response curve (35) with a maximal 1.5-4-fold increase in FRET signal in the presence of Ig1 or Ig2 targeting antibodies at 5-20 nM LYTAC (fig. S4). No significant signal increase was observed over the same concentration range using MuSK Bait. Together, these results highlight the ability of MuSK LYTAC to bind MuSK antibodies with high affinity and to co-engage these antibodies and ASGPR.

### Validation of MuSK LYTAC design with multiple MuSK MG patient samples

Having established that MuSK Bait and MuSK LYTAC bind with high affinity to four MuSK antibodies, we next tested the ability of the protein to bind a broader repertoire of antibodies found in MuSK MG patients. We acquired 11 patient serum samples (MG1-MG11) confirmed to be positive for MuSK antibodies by clinical diagnostic radioimmunoassay (RIA) (table S2). The ability of immobilized MuSK Bait to remove antibodies from the serum samples was tested using a live cell-based binding assay (CBA) and a functional AChR clustering assay to measure the pathogenic capacity of the samples (7, 9, 36). MuSK Bait was immobilized on Sepharose beads and incubated with patient serum to allow for antibody binding and depletion. Serum samples pre- and post-depletion were then tested in the CBA with HEK293 cells expressing full-length MuSK. Because we obtained a larger volume of serum from patient MG11 and limited amounts of serum from patients MG1-MG10, we conducted a series of control experiments with the MG11 sample to optimize the assay parameters. Incubation with MG11 serum resulted in a 3.5-fold increase in cell binding signal, relative to a healthy control serum sample, indicating the presence of MuSK-binding antibodies (Fig. 2A). No significant binding was detected for the MG11 serum sample or a mixture of anti-MuSK antibodies to HEK293 cells expressing GFP, indicating that the assay is selectively measuring binding to MuSK (fig. S5A). The MG11 serum was then tested after depletion with beads alone, HSA-coupled beads or MuSK Bait-coupled beads. Changes in cell binding of +2% and −30% were observed with the beads alone or HSA-coupled beads samples, respectively, whereas treatment with MuSK Bait-coupled beads resulted in an 88% decrease in signal. The weak apparent depletion by HSA-coupled beads may be caused by sample dilution or nonspecific binding. Similar degrees of depletion were observed with immobilized MuSK Bait and MuSK LYTAC, indicating that ASGPR ligand conjugation does not significantly impede antibody binding (fig. S5B). For patient samples MG1-MG10, incubation with MuSK Bait-coupled beads resulted in a significant decrease in cell binding signal in all samples except MG10, which did not show a signal above background in the pre-depletion sample (Fig. 2B). Of the remaining samples, MuSK Bait depletion of 8/9 resulted in a signal decrease to the level of the negative control sample (p>0.17).

**Figure 2.**
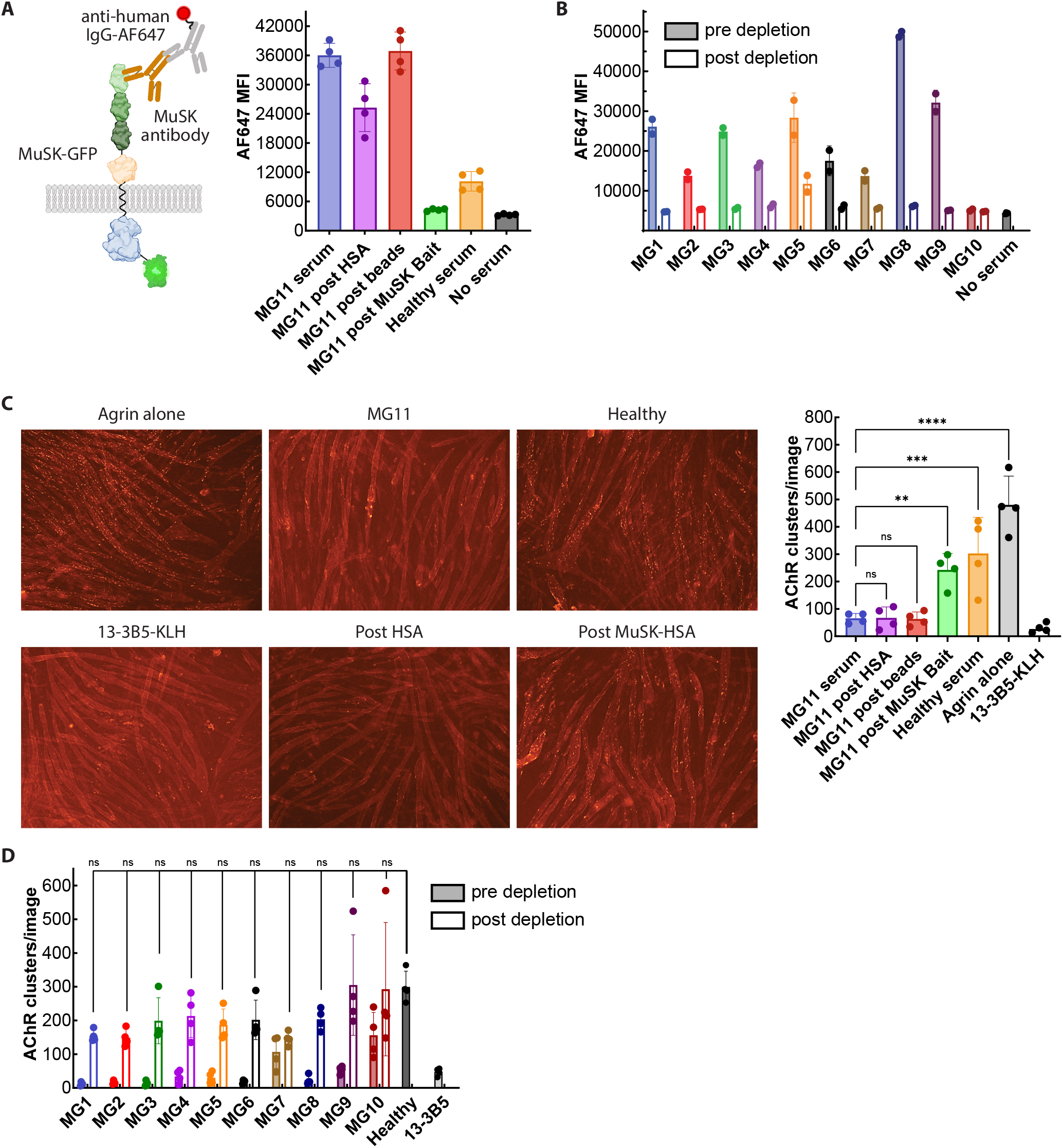
MuSK Bait binds to the majority of total and pathogenic MuSK antibodies from patients. **(A)** FACS CBA using HEK293 cells overexpressing full length MuSK-GFP with MG11 patient serum before and after incubation with MuSK Bait conjugated beads and compared to controls. Data are representative of three independent experiments. **(B)** MuSK-GFP-expressing HEK293 cell binding data with a panel of 10 MG patient samples before and after depletion by MuSK Bait-conjugated beads. Samples were analyzed in two independent experiments. **(C)** AChR clustering assays showing the number of AChR clusters per image in agrin-stimulated C2C12 mytotubes after incubation with MG11 patient serum before or after incubation with MuSK Bait-conjugated beads or controls, with representative images. Data are representative of three independent experiments. **(D)** AChR clusters per image in agrin-stimulated C2C12 mytotubes after incubation with MG patient serum before or after treatment with MuSK Bait-conjugated beads. No statistical significance for post depletion samples compared to healthy serum based on one-way ANOVA (p>0.10).

In the AChR clustering assay, MuSK antibodies with pathogenic potential block MuSK signaling, which is required for agrin-induced AChR clustering (Fig. 2C). Addition of 13-3B5-KLH led to a 93% decrease in the number of AChR clusters/image, whereas addition of healthy serum did not significantly impact the number of AChR clusters/image. In contrast to healthy serum, the addition of MG11 serum caused an 88% decrease in the AChR cluster count. MG11 serum sample incubation with beads alone or HSA-coupled beads did not lead to a significant increase in cluster count, while incubation with MuSK Bait-coupled beads led to a 3-fold increase in cluster count. As with the CBA, similar results were obtained by depletion with MuSK LYTAC instead of MuSK Bait (fig. S5C). For serum samples MG1-MG10, incubation with MuSK Bait led to an increase in cluster count as compared to the pre-treated sera for all 10 samples, though statistical significance was not reached with MG7 and MG10 samples due to the higher AChR cluster numbers of the pre-depletion samples (Fig. 2D). The post-depletion cluster numbers for all 10 samples were not statistically different than the healthy serum-treated samples (p>0.10). Overall, results from two orthogonal cell-based assays demonstrate the ability of MuSK Bait to bind the majority of total and pathogenic MuSK antibodies from multiple patient samples.

### In vitro activity and mechanism of MuSK LYTAC

We tested the ability of MuSK LYTAC to mediate ASGPR-dependent uptake and degradation of MuSK antibodies in a series of cell-based experiments using the Hep G2 human liver carcinoma cell line. In the first set of experiments, we measured the ability of MuSK LYTAC to mediate cellular binding and uptake of AF647-labeled MuSK antibodies in Hep G2 and ASGPR 1/2 double knockout Hep G2 cells (Hep G2 *ASGPR*^−/−^). MuSK LYTAC induced concentration-dependent uptake of MuSK Ig1 or Ig2 domain-specific antibodies in Hep G2 cells (Fig. 3A). Minimal uptake was observed with MuSK antibody alone, with AChR antibodies (mAb35 and mAb637) or with MuSK Bait conjugated with linker-ligands containing non-binding, ASGPR ligand enantiomers (MuSK LYTAC-ent) (Fig.3A, fig. S6). MuSK LYTAC did not mediate antibody uptake in Hep G2 *ASGPR*^−/−^ cells (Fig. 3A). To test the translatability of the observed activity, we conducted antibody uptake studies in primary human, rat, canine and cynomolgus monkey hepatocytes. Unlike MuSK Bait, MuSK LYTAC mediated antibody uptake in all primary hepatocytes tested, when compared to the antibody alone (fig. S7). Similar studies were conducted with MuSK antibodies labeled with pHrodo green, which is weakly fluorescent at neutral pH and becomes increasingly fluorescent under acidic conditions. Dose-dependent increases in signal were observed with pHrodo green-labeled MuSK1A, 13-3B5 or 13-3B5-KLH in the presence of MuSK LYTAC (fig. S8). Minimal signal change was observed with MuSK antibody alone, antibody plus MuSK Bait, or with MuSK LYTAC in *ASGPR*^−/−^ cells. These results are consistent with the ability of MuSK LYTAC to deliver antibodies to an acidic environment within the cell.

**Figure 3.**
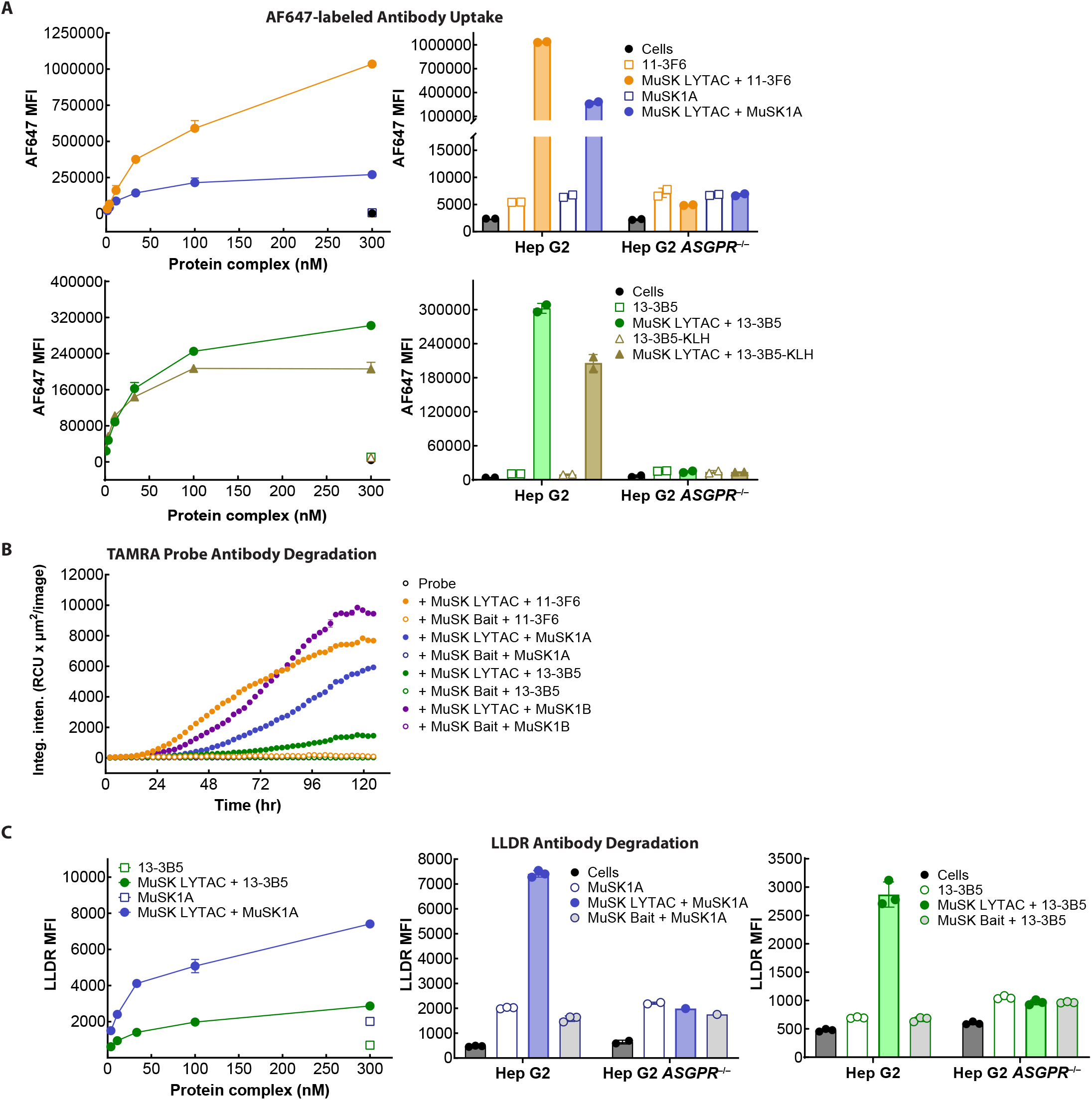
LYTAC-mediated, dose-dependent antibody uptake and degradation in Hep G2 cells. **(A)** AF647-labeled MuSK antibody uptake studies with 11-3F6, MuSK1A (top left), 13-3B5 and 13-3B5-KLH (bottom left) with AF647 mean fluorescence intensity (MFI) measured by flow cytometry plotted as a function of protein concentration (antibody or 1:1 antibody + MuSK LYTAC). AF647 signal with cells alone or with 300 nM protein (antibody or 1:1 antibody + MuSK LYTAC) in WT Hep G2 or Hep G2 *ASGPR* knockout (*ASGPR*^−/−^) cells (top and bottom right) **(B)** MuSK antibody degradation using a TAMRA/QSY7-conjugated anti-human F(ab’)2. Intensity measured by microscopy is shown as a function of time. Probe mixed with MuSK Bait or MuSK LYTAC and the indicated antibodies at a 1:1:1 ratio (50 nM each). **(C)** MuSK antibody degradation measured with Lysolight DR (LLDR)-labeled antibodies. Concentration-dependent signal increase with MuSK LYTAC and LLDR-labeled 13-3B5 or MuSK1A (left). Signal with cells alone or with 300 nM LLDR-labeled antibody or antibody complex in Hep G2 or Hep G2 *ASGPR*^−/−^ cells (center, right). Data are representative of three independent experiments.

LYTAC-induced antibody degradation was measured using two different methods. First, we used a TAMRA-QSY7 fluorophore-quencher pair-labeled anti-human IgG F(ab′)2 that generates an increase in fluorescence upon cellular internalization and protein degradation (37). As shown in Figure 3B, the addition of probe to MuSK LYTAC and Ig1- or Ig2-binding MuSK antibodies to Hep G2 cells led to a steady increase in fluorescence signal that plateaus after 4-5 days, as measured using time-lapsed microscopy. Degradation was also observed with monovalent Ig1- or Ig2-binding antibodies. Minimal antibody degradation was observed with MuSK Bait treatment (fig. S9).

Antibody degradation was also measured using antibodies directly labeled with LysoLight Deep Red (LLDR), which only fluoresces upon cleavage by the lysosomal protease Cathepsin B (38). LLDR-labeled 13-3B5 or MuSK1A were mixed at a 1:1 molar ratio with MuSK LYTAC and incubated with Hep G2 cells for 2 hr before measuring the fluorescence signal by FACS to monitor degradation. MuSK LYTAC mediated a dose-dependent increase in degradation over the 3-300 nM concentration range. At 300 nM, 4.1- and 3.8-fold signal increases vs. antibody alone were observed for 13-3B5 and MuSK1A, respectively (Fig. 3C). As with the quenched TAMRA probe method, antibody degradation was LYTAC- and ASGPR-dependent, as minimal degradation was observed on treatment with MuSK Bait or with MuSK-LYTAC in Hep G2 *ASGPR*^−/−^ cells. Together, the results from these cellular assays demonstrate the ability of MuSK LYTAC to drive uptake and degradation of Ig1- and Ig2-domain binding MuSK antibodies via ASGPR.

### LYTAC-mediated clearance of MuSK antibodies in vivo

We next tested MuSK LYTAC activity in rodents. First, the serum and tissue pharmacokinetics (PK) of MuSK LYTAC were investigated to better understand clearance properties imparted by the ASGPR ligands. Rats were administered MuSK LYTAC intravenously (IV) or subcutaneously (SC), or MuSK Bait SC and serum levels were assessed using sandwich ELISA methods to measure total (with and without ASGPR ligand) or conjugated (with ASGPR ligand) MuSK-HSA. The two assays enabled tracking of low levels of unconjugated protein in the starting material or deconjugation that occurred during the experiment. IV or SC administration of MuSK LYTAC resulted in rapid elimination and low serum exposure of the molecule (fig. S10, A and B). In comparison, MuSK Bait administered SC demonstrated substantially higher serum exposure, and a half-life consistent with other HSA fusions in rat (39). Despite the low serum exposure, MuSK LYTAC was detected in the liver but not spleen or kidney at 2 hr post dose and was undetectable by 24 or 168 hr, depending on the dose level (fig. S10, C and D). The combination of the serum and tissue PK results suggests that the rapid clearance and low apparent MuSK LYTAC bioavailability is due to rapid on-target clearance through the liver, which is the main site of ASGPR expression. Similar serum PK profile differences of SC administered MuSK LYTAC and MuSK Bait were observed in mice (fig. S11). Having demonstrated that MuSK LYTAC displays rapid serum clearance and liver targeting in rodents, we next tested the ability of MuSK LYTAC to clear pathogenic MuSK antibodies in vivo. C57BL/6 mice were administered biotinylated 13-3B5 or MuSK1A IV, followed by SC treatment with MuSK LYTAC or controls 16 hr later. Serum samples were collected over the next 24-48 hr and analyzed for total biotin-MuSK antibody levels (Fig. 4A). MuSK LYTAC mediated dose-dependent clearance of both MuSK1A and 13-3B5. (Fig. 4B and fig. S12 A, B and C). Four hr after treatment, 10 mg/kg MuSK LYTAC promoted decreased levels of MuSK1A and 13-3B5 by 84% and 99% respectively compared to the pre-treatment (t=0 hr) levels and these decreases were significantly greater than those observed in the untreated groups. Activity measured by area under the curve (AUC) or % clearance at 4 hr, was dependent on the initial antibody dose (Fig. 4C). In contrast to MuSK LYTAC, 10 mg/kg MuSK LYTAC-ent treatment led to a slight stabilization of MuSK1A levels at 4 hr that persisted through 48 hr (Fig. 4B). The inability of MuSK LYTAC-ent to clear MuSK antibodies is consistent with the lack of activity in cellular uptake assays.

**Figure 4.**
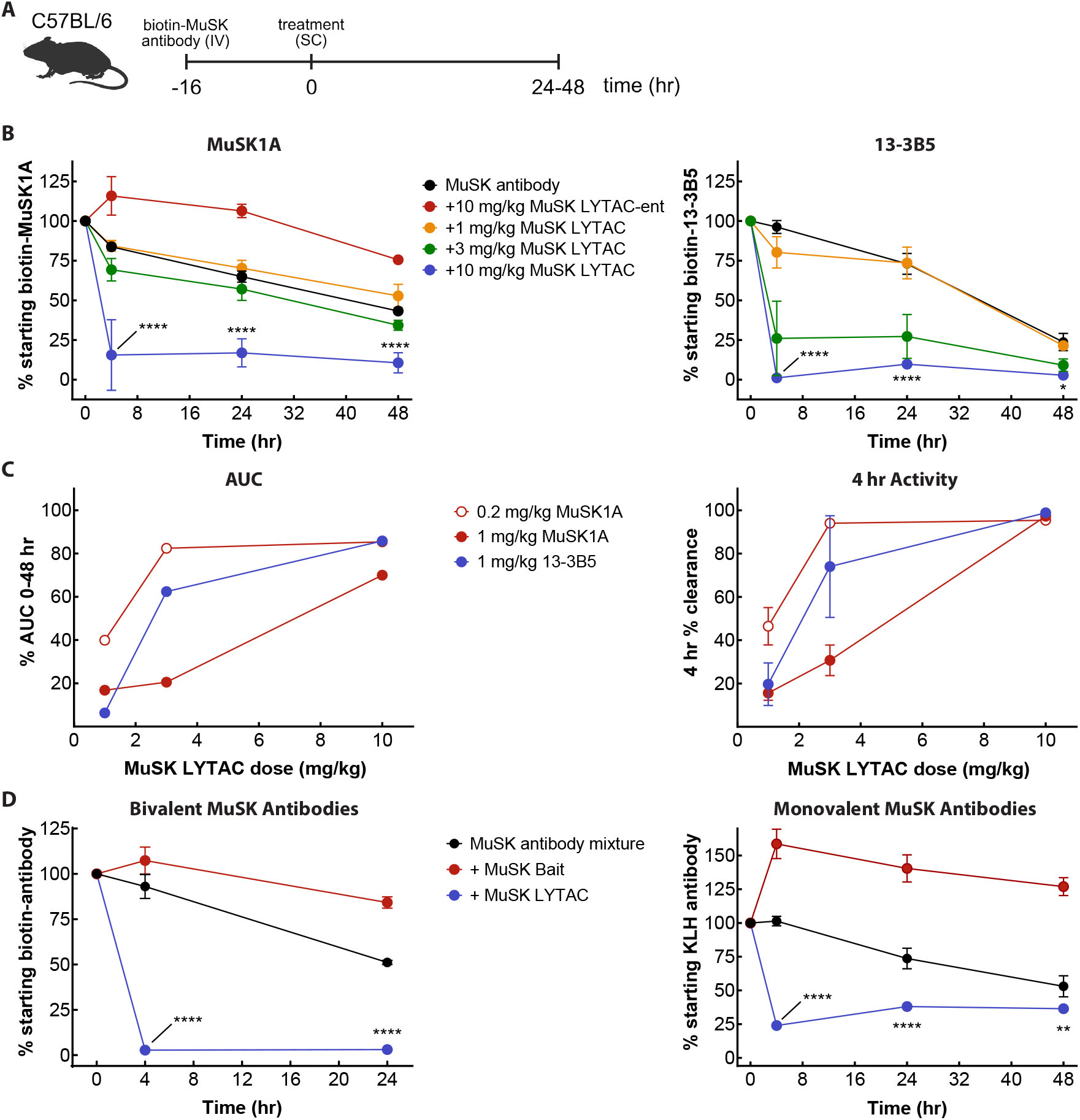
MuSK LYTAC clears Ig1- and Ig2-binding MuSK antibodies in a dose-dependent manner. **(A)** Schematic of MuSK antibody clearance studies in C57BL/6 mice injected with biotinylated MuSK antibody followed by treatment at t=0. **(B)** With 1 mg/kg MuSK antibody, percent of pre-treatment (t=0) serum MuSK1A levels (left) or 13-3B5 levels (right) after treatment with the indicated dose range of MuSK LYTAC or with 10 mg/kg MuSK LYTAC-ent. **(C)** MuSK antibody clearance based on area under the curve (AUC) 0-48 hr (left) or % clearance at 4 hr (right) as a function of MuSK LYTAC dose. **(D)** Percent of pre-treatment (t=0) MuSK antibody levels after dosing with 10 mg/kg MuSK LYTAC or 10 mg/kg MuSK Bait with 1 mg/kg (total) mixture of three bivalent (left) or monovalent (right) MuSK antibodies (13-3B5, 11-3F6, MuSK1A). Statistics: 2way ANOVA, comparison of 10 mg/kg MuSK LYTAC to MuSK antibody or MuSK antibody mixture,**** = P < 0.0001, ** = P < 0.005; n = 3 animals/group. Data are representative of two independent experiments.

MuSK MG is known to be caused by a polyclonal population of autoantibodies, so we tested the ability of MuSK LYTAC to clear mixtures of monoclonal antibodies. Mixtures (1:1:1, 1 mg/kg total) of bivalent, biotinylated 13-3B5, 11-3F6 and MuSK1A or monovalent 13-3B5-KLH, 11-3F6-

KLH and MuSK1A-KLH were administered to mice followed by treatment with 10 mg/kg MuSK LYTAC or MuSK Bait. For the bivalent mixture, MuSK LYTAC treatment decreased antibody levels by 97% relative to t=0, compared to a 7% decrease in the untreated group and a 7% increase in the MuSK Bait-treated group (Fig.4D and fig. S12, D and E). Similar trends occurred with the monovalent antibody mixture, with 76% depletion at 4 hr relative to pre-treatment levels in the MuSK LYTAC group.

To better understand the mechanism of antibody clearance, we expanded our pharmacodynamic studies to include ELISA measurements of MuSK antibody and LYTAC levels in liver, kidney and spleen. Biotin-MuSK1A was injected, followed by MuSK LYTAC or MuSK Bait 24 hr later (Fig. 5A). MuSK LYTAC caused rapid clearance of MuSK1A from the serum, as demonstrated in the above studies, while MuSK Bait induced MuSK1A stabilization, when compared to the untreated group (MuSK1A alone) (Fig. 5B). Importantly, by 30 minutes post treatment, MuSK LYTAC treatment led to appreciably higher MuSK1A levels in the liver than MuSK Bait treatment or no treatment, and this difference persisted through 24 hr (Fig. 5C). By 72 hr, MuS-K1A was no longer detected in the liver of LYTAC-treated mice. MuSK1A was not detected in the spleen or kidney for any group. As with the above-described PK experiments, measurement of MuSK Bait and MuSK LYTAC levels further supports targeting of MuSK LYTAC to the liver. Like MuSK1A, MuSK LYTAC was detected in the liver by 30 minutes and levels peaked at 2 hr before declining below the lower limit of detection by 72 hr. In contrast to MuSK Bait, no detectable MuSK LYTAC was present in spleen or kidney. Overall, the results demonstrate that MuSK LYTAC binds to MuSK antibodies in circulation and delivers them to the liver for degradation.

**Figure 5.**
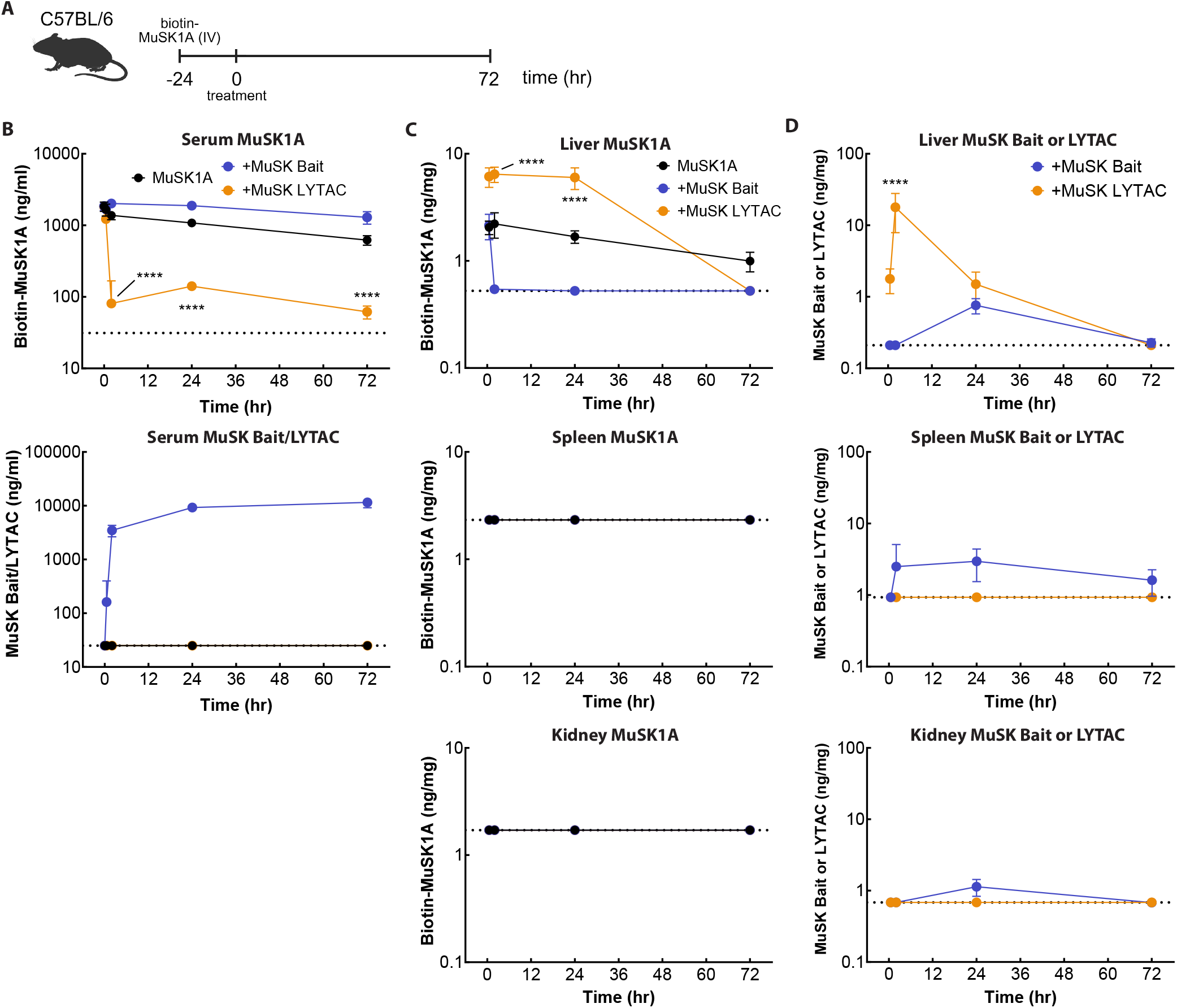
MuSK LYTAC clears MuSK1A from the serum and causes transient MuSK1A accumulation in the liver of mouse. **(A)** Schematic of MuSK antibody clearance study in C57BL/6 mice injected with 1 mg/kg biotin-MuSK1A followed by treatment with 10 mg/kg MuSK Bait or 10 mg/kg MuSK LYTAC with tissue and serum measurements of antibody and MuSK LYTAC or MuSK Bait. **(B)** Serum levels of biotin-MuSK1A (top) and MuSK Bait or MuSK LYTAC (middle). **(C)** Levels of biotin-MuSK1A in liver (top), spleen (middle) and kidney (bottom). **(D)** Levels of MuSK Bait or MuSK LYTAC in liver (top), spleen (middle) and kidney (bottom). Dotted lines indicate ELISA limits of detection. Statistics: 2way ANOVA, comparison of MuSK LYTAC to MuSK1A (B and C) or MuSK LYTAC to MuSK Bait (D), **** = P < 0.0001. Data are representative of two independent experiments with n = 3 animals/timepoint >0 hr.

### MuSK LYTAC activity in a mouse model of MG

Previous reports have described a passive immunization MG disease model through injection of the monovalent MuSK Ig1-binding antibodies 13-3B5 and 11-3F6 into NOD/SCID mice (13). We established a similar disease model using a single intraperitoneal (IP) injection of 1.5 mg/kg 13-3B5-KLH to test the in vivo activity of MuSK LYTAC (Fig. 6A). Passive immunization with 13-3B5-KLH led to maximal weight loss and grip strength decreases on day 11, with a 7.9% average weight decrease, and a 59% grip strength decline (Fig. 6B). These decreases were prevented by administration of the FcRn inhibitor efgartigimod on day 3, which led to a 71% decrease in 13-3B5-KLH levels on day 7. The efgartigimod dose and route of administration were based on previously reported work (40). In this model, subcutaneous (SC) injection of MuSK-LYTAC on day 3 also prevented weight loss and grip strength decline, with values indistinguishable from naïve mice on day 11 (Fig. 6C). MuSK LYTAC treatment led to a 90% decrease in 13-3B5-KLH antibody levels, relative to the untreated group, four hours after treatment. This decrease in 13-3B5-KLH was maintained on day 7; 13-3B5-KLH levels were 83% lower in MuSK LYTAC group than in the untreated animals. To compare selectivity profiles of MuSK LYTAC and efgartigimod for IgG, we conducted a single dose pharmacodynamic study in C57BL/6 mice, which unlike NOD/SCID mice, produce IgG (Fig. 6D). As in the MG disease model, a single dose of 13-3B5-KLH was administered, followed by 25 mg/kg efgartigimod or 10 mg/kg MuSK LYTAC. Four hours after treatment, MuSK LYTAC and efgartigimod reduced 13-3B5 levels by 98% and 46%, respectively, compared to a 4% decrease in the untreated group (Fig 6E). By 24 hr, 13-3B5-KLH levels in the MuSK LYTAC, efgartigimod and untreated groups decreased by 88%, 74% and 28%, respectively. In contrast, MuSK LYTAC treatment did not lead to significant changes in total mouse IgG levels, while efgartigimod treatment caused a sustained ∼60% decrease in total mouse IgG. This result highlights the mechanistic differences of the antigen-specific LYTAC approach. We next sought to establish a similar passive immunization model with the Ig2 domain binder MuSK1A to test MuSK LYTAC activity. Three injections of 0.5 or 2.5 mg/ kg monovalent MuSK1A-KLH induced a decrease in body weight and grip strength in NOD/SCID mice to a similar extent as three injections of 2.5 mg/kg Ig1 domain binder 13-3B5-KLH (fig. S13). As seen with Ig1-binding antibodies, monovalency enhances pathogenicity, as three doses of up to 10 mg/kg bivalent MuSK1A do not cause significant changes to body weight or grip strength over 10 days (fig. S14). LYTAC activity was tested after administering a single injection of MuSK1A-KLH (Fig. 7A). Mice were administered a single treatment of MuSK LYTAC on either day 1, day 3 or day 6 after MuSK1A-KLH injection. Mice injected with MuSK1A-KLH showed a measurable decrease in body weight by day 7 and continued to decline through day 10, at which time mice were euthanized (Fig. 7, B and C). On day 10, these animals also showed a 69% average decrease in grip strength. MuSK LYTAC administration on days 1 or 3 largely prevented the decline in body weight and grip strength, while administration on day 6 led to intermediate improvements in body weight relative to the untreated animals. On day 17, the body weights and grip strengths of animals in all of the MuSK LYTAC-treated groups were indistinguishable from the naïve animals, regardless of the day MuSK LYTAC was administered (Fig 7D). Together, the disease model data demonstrate the ability of MuSK LYTAC to inhibit development of disease symptoms through selective clearance of MuSK antibodies.

**Figure 6.**
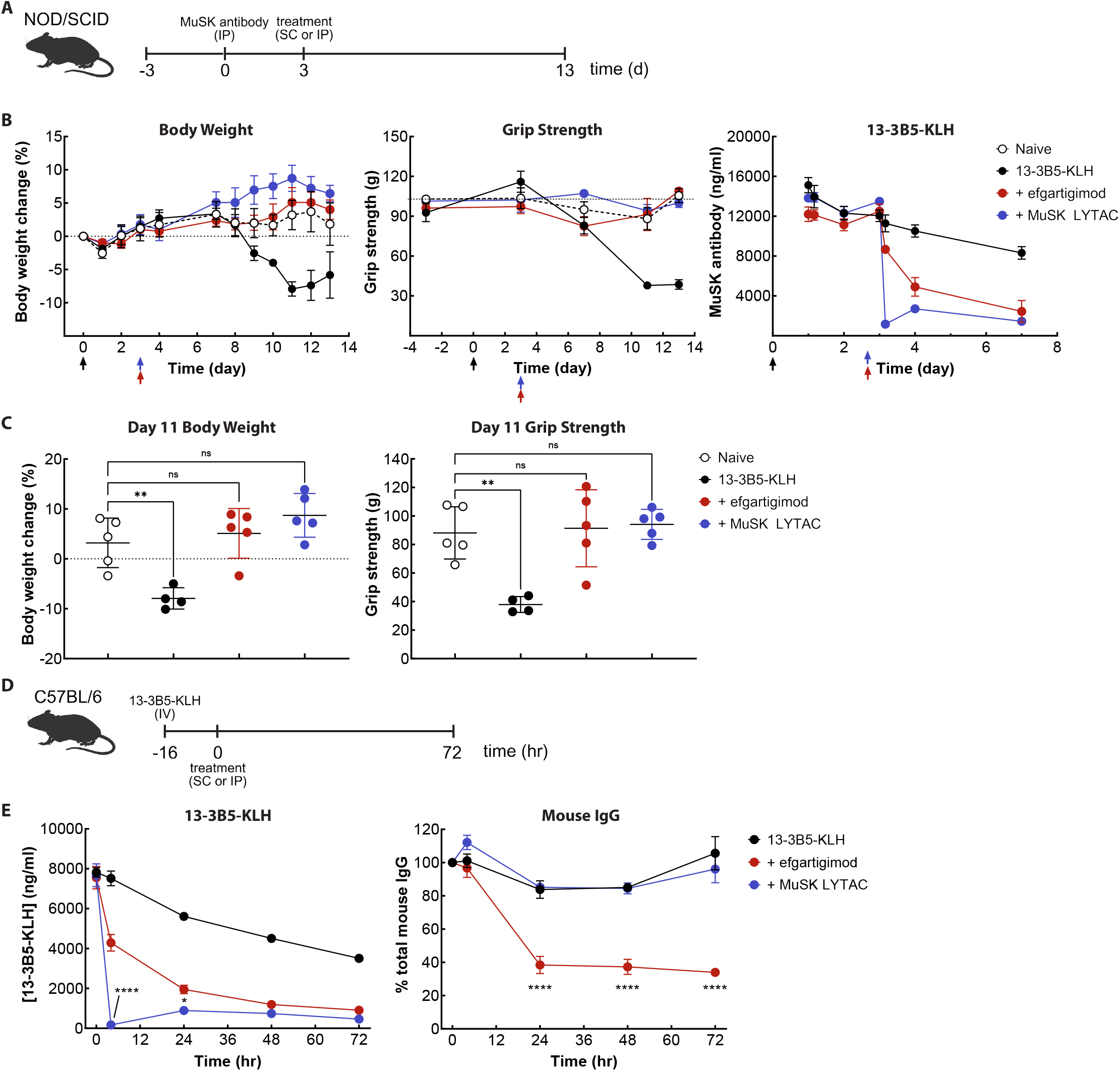
MuSK LYTAC prevents disease onset in the passive transfer model of MuSK-MG and does not deplete total IgG. **(A)** Schematic of passive transfer MuSK-MG mouse model induced by 1.5 mg/kg 13-3B5-KLH injection on day 0, followed by treatment with 25 mg/kg efgartigimod IP or 10 mg/kg MuSK LYTAC SC on day 3. **(B)** Body weight (left), grip strength (middle), and MuSK antibody level (right) changes measured over the course of the study. **(C)** Body weight (left) and grip strength (right) comparisons on day 11. One way ANOVA P value: ** <0.01. Data are representative of two independent experiments with n = 4 (13-3B5-KLH group) or 5 animals (other groups) per group. **(D)** Schematic of MuSK antibody clearance study with 25 mg/kg efgartigimod IP or 10 mg/kg MuSK LYTAC SC treatment 16 hr post 1 mg/kg 13-3B5-KLH dose. **(E)** 13-3B5 levels (left) and total mouse IgG levels (right) prior to treatment through 72 hr post treatment. Statistics: 2way ANOVA, comparison of MuSK LYTAC to efgartigimod, **** = P < 0.0001, * = P < 0.05; n = 3 animals/group.

**Figure 7.**
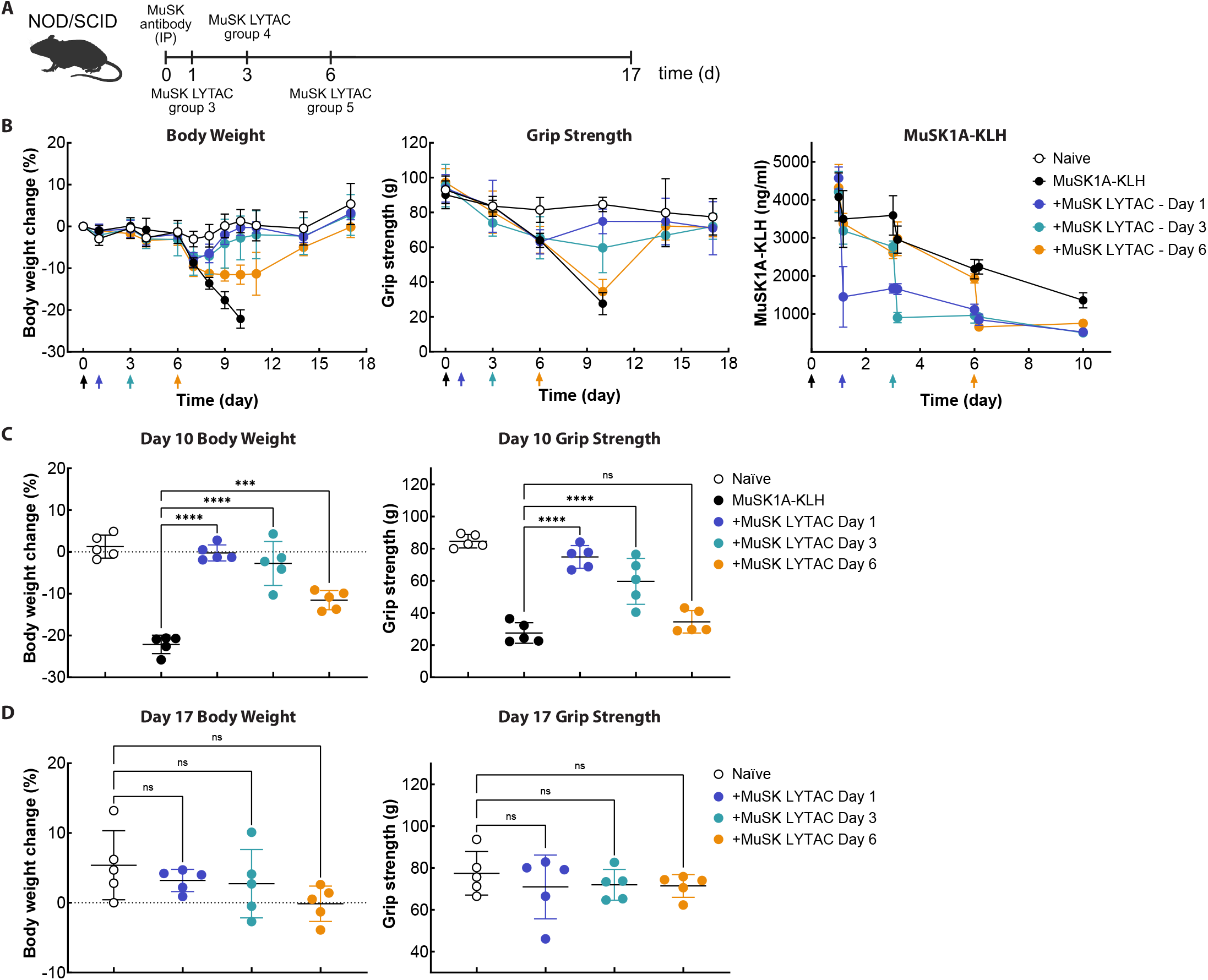
MuSK LYTAC prevents disease onset in delayed dose MuSK1A MG-mouse model. **(A)** Schematic of passive transfer MuSK-MG mouse model with 10 mg/kg MuSK LYTAC treatment on day 1, 3, or 6. **(B)** Body weight (left), grip strength (middle), and serum MuSK antibody levels (right) measured after 1.5 mg/kg MuSK1A-KLH injection on day 0 with MuSK LYTAC treatment on day 1, 3 or 6 (arrows). **(C)** Body weight (left) and grip strength (right) comparisons on day 10. One way ANOVA P value: *** <0.01. Data are representative of 2 independent experiments with n = 5 animals/ group.

## Discussion

LYTACs have been proposed as therapeutics for the elimination of circulating proteins that are drivers of disease. Autoimmune disorders such as MuSK MG are an enticing application of LYTAC technology because autoantibodies against extracellular self-antigens directly cause disease. These disorders are often managed with generalized immunosuppressive agents that address inflammation (e.g., corticosteroids), reduce total IgG (e.g., FcRn antagonists or plasmapheresis) or deplete B cells (e.g., rituximab). These non-specific therapies can have significant systemic side effects and render patients less responsive to vaccines and more susceptible to infection (41–43). Instead, we sought to specifically target disease-driving autoantibodies by combining the rapid degradation potential of LYTAC technology, which utilizes the high-capacity scavenging receptor ASGPR, with a bait construct comprising the disease-relevant domains of MuSK linked to human serum albumin. We hypothesized that this precision immunology approach would result in the rapid and selective removal of autoantibodies that drive MuSK MG without broadly compromising immune system function. MuSK Bait was rationally designed to include both the Ig1 and Ig2 domains of the native MuSK protein, which are the targets of the majority of autoantibodies in MuSK MG patients (7, 27, 29). While the pathogenicity of Ig1 binders was previously established, our results demonstrate that patient-derived Ig2 binders are also capable of driving an MG phenotype in vivo – confirming that effective treatment of MuSK MG will require removal of Abs that bind to either Ig1 or Ig2. MuSK Bait was able to capture all detectable MuSK-binding antibodies present in the serum of MuSK MG patients and rendered those samples incapable of disrupting AChR clusters in C2C12 myotubes. Importantly, addition of ASGPR ligands to create MuSK LYTAC does not significantly impair binding to MuSK antibodies.

MuSK LYTAC is pharmacologically active, capable of degrading MuSK antibodies via ASGPR-dependent internalization and trafficking to the lysosome in either Hep G2 cells or primary human hepatocytes. Mouse pharmacodynamic models demonstrate the plausible in vivo translatability of this mechanism, showing that subcutaneously administered MuSK LYTAC efficiently directs circulating MuSK antibodies to the liver for degradation. In a murine passive immunization model where patient-derived MuSK antibodies induce weight loss and muscle weakness consistent with MuSK MG patient symptoms, a single dose of MuSK LYTAC was sufficient to prevent disease onset. In contrast to efgartigimod, MuSK LYTAC also achieved efficacy with the expected precision, selectively eliminating MuSK antibodies while preserving overall IgG concentration at levels equivalent to the untreated group.

The ABLate mechanism of action underlying MuSK LYTAC offers multiple significant advantages compared to alternative modalities proposed for the treatment of antibody-driven autoimmune diseases. First, the use of a “bait” to enable elimination of only disease-causing antibodies, while leaving the remainder of the immune repertoire intact, stands in contrast to indiscriminate removal all B cells or reduction of all IgG. Second, the depth of depletion achieved by MuSK LYTAC offers better potential for pharmacologic effect compared to FcRn inhibitors, which only offer partial removal of IgG and therefore leave a subset of pathogenic autoantibodies, including those of the IgA isotype, unaddressed (44, 45). Third, the ABLate approach leverages the high-capacity, liver-specific ASGPR – a receptor naturally designed to clear glycoproteins from circulation – enabling more rapid and deeper depletion of autoantibodies compared to other antigen-specific modalities that rely on less efficient cell surface receptors for internalization (46, 47). A similar approach clearing beta-1 adrenergic receptor antibodies with ASGPR has been reported (48). Finally, the event-based pharmacology of LYTACs – wherein transient interaction of drug and autoantibodies is sufficient to eliminate the cause of disease – offers benefits versus traditional occupancy-based pharmacology, such as “decoy” antigen approaches or recently described MuSK agonists, where the need for high stoichiometry between drug and target can lead to durability and dosing challenges (49–51). Conversely, LYTACs have the advantage of irreversible removal of a pathogenic autoantibody target that would not induce immune complex formation. The efficacy of therapeutic MuSK agonists also depends on the specific repertoire of MuSK antibodies in each patient, with the presence of “naturally occurring” MuSK agonist Abs proving particularly problematic (52). The mechanism for MuSK LYTAC, in contrast, is agnostic to antibody repertoire and should therefore be effective across all patients.

There are potential limitations to clinical application of MuSK LYTAC. Epitope spreading has been reported to occur in 19% of MuSK MG patients (27), and MuSK antibodies that bind to either the Ig3 or Fzd domain would not be removed with MuSK LYTAC. The risk associated with such epitope spreading is expected to be minor due to the relative paucity of reported pathogenic MuSK antibodies that bind outside of Ig1 and Ig2 (7, 28, 29). Dosing levels and frequency represent another potential challenge in the context of patients that are continually producing polyclonal MuSK antibodies, as opposed to discreet target administration of select monoclonal target antibodies in mouse pharmacodynamic models. There is evidence from MG clinical immunoadsorption studies, however, that even incomplete pulsatile depletion of autoantibodies maintains suppression below baseline for up to 2-3 weeks following the withdrawal of therapy, due to the relatively low fractional synthesis rate of IgG (53). Potential immunogenicity induced through the administration of a drug containing self-antigen, against which MuSK MG patients are already sensitized, represents an additional issue to consider before application as a therapeutic. Immunogenicity should be limited due to the rapid clearance of MuSK LYTAC to hepatocytes in the liver, thus limiting the opportunity for interaction with antigen-presenting cells or B cells present in lymphoid tissue. Hepatocytes have also been described as hubs of tolerance induction, which may further limit the immune-stimulating potential of MuSK LYTAC (54).

The LYTAC-based approach to depletion of disease-driving antibodies has broad applicability that extends beyond MuSK MG. All autoimmune diseases that are driven by the production of antibodies against a known target protein could be addressed using the same strategy. Careful “Bait” engineering would be required for each new disease to ensure both structural integrity and ability to capture all pathogenic autoantibodies, but patients suffering from autoantibody-driven diseases such as AChR MG, Graves’ disease, neuromyelitis optica spectrum disorder, thrombotic thrombocytopenic purpura, myelin oligodendrocyte glycoprotein antibody-associated disease and membranous nephropathy may benefit from this modality. Successful translation of this concept to the clinic for one autoantibody-driven disease would provide proof of concept for other diseases within this category, helping shift the standard of care away from generalized immunosuppression and targeted non-selective therapy towards true precision elimination of disease mediators.

## Materials and Methods

### Study design

The purpose of this work was to design, generate and evaluate a LYTAC that selectively binds and degrades pathogenic MuSK autoantibodies that drive MG. A recombinant MuSK-HSA fusion protein was produced and tested for binding to MuSK antibodies from patient samples or produced recombinantly. A panel of 11 patient samples and four different recombinant antibodies were used to test the universality of the construct. MuSK LYTAC was generated by conjugating the fusion protein with ASGPR ligands connected by a flexible linker and tested is a series of in vitro and in vivo activity studies. Antibody uptake and degradation were evaluated in Hep G2 cells and in some cases primary hepatocytes, given the high expression of ASGPR, using fluorophore-labeled antibodies. Both monovalent and bivalent antibodies were tested due to the high propensity of monovalent IgG4 autoantibodies in MuSK-MG. Serum and tissue PK were measured in mice and rats, and activity studies were conducted in mice. ELISA methods were used to detect the ability of MuSK LYTAC to clear MuSK antibodies from circulation and drive accumulation in the liver, spleen or kidney. Finally, we evaluated MuSK LYTAC in a mouse MuSK-MG disease model caused by monovalent MuSK antibodies by monitoring body weight, grip strength and MuSK antibody levels. The sample sizes for in vivo studies were determined based on previous experiments. The studies were not blinded. Statistical methods to determine data significance and the number of repeats conducted for each study are indicated in the appropriate figure legends.

### Protein production, purification and conjugation

Sequences for all recombinant antibodies used in this study were sourced from public databases. MuSK-specific antibodies 13-3B5 and 11-3F6 were obtained from WO2020055240A1; MuSK1A and MuSK1B were obtained from protein data bank (https://www.rcsb.org/) from accession codes 6YWR and 6WYT. The sequence for AChR-specific antibody mAb35 was obtained from protein data bank (https://www.rcsb.org/) accession code 5HBV and the mAb637 sequence was from (55). All recombinant antibody variable regions (vL and vH) were cloned into a human IgG1 framework containing wildtype residues or L234A/L235A. For production of monovalent autoantibodies, we utilized a duoBody controlled Fab arm exchange process in our IgG1 constructs in which one half of an antibody contains a F405L point mutation and the pairing side contains a K409R mutation.

### MuSK Bait production and purification

MuSK-HSA (MuSK Bait) protein was expressed in ExpiCHO cells using the ExpiFectamine™ CHO Transfection Kit (Thermo Fisher), according to the manufacturer’s instructions. Following protein expression, the protein was purified on a CaptureSelect™ Human Albumin Affinity Matrix column (Thermo Fisher), followed by anion exchange chromatography on a Q-HP column (Cytiva). In the first purification step, protein was loaded onto the column, washed with 3 column volumes (CV) of 1x phosphate buffered saline (PBS) (pH 7.5) and eluted using 3CV of 0.1 M glycine pH 3.2. Protein was then neutralized to pH 8 and diluted threefold with 20 mM Tris pH 8. In the second purification step, protein was loaded onto the column and sequentially washed with 3CV of 20 mM Tris pH 8, 3CV of 20 mM Tris pH 8 containing 25 µg/mL N-acetylcysteine, and 3CV of 20 mM Tris pH 8. The protein was then eluted using a 20CV linear gradient of 1M sodium chloride. Fractions containing the purified protein were collected and assessed using intact mass analysis for mass confirmation, SEC-HPLC (>95% main peak), SDS-PAGE for purity, and Endosafe® nexgen-PTS™ for endotoxin level.

### Antibody production purification and monovalent antibody assembly

Antibodies were expressed in Expi-293 cells using the Expi293™ Expression System Kit (Thermo Fisher),according to the manufacturer’s instructions. Following protein expression, the antibody was purified on a MabSelect SuRe affinity column (Cytiva), followed by cation exchange chromatography on a SP-HP column (Cytiva). In the first purification step, the antibody was loaded onto the column, washed with 5CV of 1x PBS (pH 7.5), and eluted using 5CV of 0.1 M acetic acid pH 3.5. The antibody was then neutralized to pH 5.2 and diluted threefold with 30 mM sodium acetate pH 5.2. In the second purification step, the antibody was loaded onto the column, washed with 30 mM sodium acetate pH 5.2, and eluted using a linear gradient of 1 M sodium chloride. Fractions containing purified antibody were pooled and buffer exchanged into 1x PBS (pH 7.4) using a 40k Zeba Desalting Spin column (Thermo Fisher). The purified antibody was assessed using intact mass analysis for mass confirmation, SEC-HPLC (>95% main peak), SDS-PAGE for purity, and Endosafe® nexgen-PTS™ for endotoxin level.

Monovalent antibodies were generated using a controlled Fab-arm exchange process (cFAE) (56). Parental antibodies were mixed at a 1:1 molar ratio, adjusted to 1 mg/ mL in 1x PBS (pH 7.4), and reduced with a final concentration of 25 mM 2-mercaptoethylamine•HCl. Samples were incubated for 3 hours at 22 °C, and reduction efficiency was confirmed using SDS-PAGE. The reducing agent was removed using 40k Zeba Desalting Spin columns pre-equilibrated in 1x PBS (pH 7.4), and the samples were allowed to re-oxidize overnight at 4 °C. The controlled Fab-arm exchange reaction was assessed using intact mass analysis to confirm reassembled monovalent antibody, SEC-HPLC (>95% main peak), and SDS-PAGE for purity.

### MuSK Bait conjugation

Following purification, MuSK Bait was concentrated to 40 mg/mL using Amicon Ultra Centrifugal Filters (Millipore Sigma). The protein was buffer exchanged into BBSE, Ph 8.3 buffer (100 mM Borate, 50 mM NaCl, 1 mM EDTA) using Zeba Desalting Columns. MuSK Bait was mixed with 8 molar equivalents of linker-ASGPR ligand containing a pentafluorophenyl (PFP) ester for conjugation to lysines through amine-reactive chemistry. Prior to the reaction, the linker-ASGPR ligand was reconstituted to 30 mM in dimethylacetamide (DMA). The reaction proceeded for 16 hr at 30 °C before unreacted linker-ASGPR ligand was removed by size exclusion chromatography (Cytiva) and buffer exchanged into PBS, pH 7.4. Purified MuSK LYTAC was sterile filtered with 0.22 µm PES syringe filters. The final concentration of MuSK LYTAC was measured using a Nanodrop spectrophotometer. The purified conjugate was assessed using SEC-HPLC (95% main peak) and SDS-PAGE for purity. Hydrophilic interaction liquid chromatography (HILIC) and intact mass analysis (Waters Xevo-TQ-S) were used to estimate the average number of linker-ASGPR ligands per MuSK Bait (conjugation ratio) (Fig. S1).

### MuSK antibody dye labeling

MuSK antibodies were conjugated with fluorescent dyes for assessment of LYTAC efficacy in cellular assays. Briefly, AF-647 containing an amine-reactive handle was conjugated to MuSK antibodies using Alexa Fluor Fluorescent Protein Labeling Kits (Thermo Fisher, A20173). Amine-reactive pH-sensitive pHrodo Green, STP Ester dye (Thermo Fisher, P36012) was conjugated to the antibodies according to the manufacturer’s protocol. Lastly, LysoLight™ Deep Red SDP ester was conjugated to the antibodies, as a tool for visualizing the lysosomal localization and degradation, using LysoLight™ Deep Red Antibody Labeling Kits (Thermo Fisher, L36002). The final concentration and degree of labeling of the dye labelled antibodies was determined by Nanodrop and the monomer purity was measured with SEC-HPLC.

### MuSK LYTAC LC-MS peptide mapping

200 µg of purified MuSK LYTAC was denatured in 6 M Guanidine HCl, 250 mM Tris, and reduced with tris(2-carboxyethyl)phosphine (at a final concentration of 5 mM) for 30 minutes at 37 °C. Once denatured and reduced, the protein was alkylated with iodoacetamide (at a final concentration of 10 mM) for 45 minutes at 22 °C in the dark. The protein was then buffer exchanged into 50 mM ammonium bicarbonate for enzymatic digestion. The protein was digested with Glu-C at a ratio of 1:20 (enzyme:protein) overnight at 37 °C. Glu-C was selected over trypsin to minimize missed cleavages, due to the lysine conjugation used for attachment of the linker-ligand. The reaction was quenched with 0.2% formic acid, diluting the sample to a final concentration of 0.5 mg/mL for subsequent LC-MS analysis.

The MuSK LYTAC digest was analyzed by a Waters ACQUITY Premier UPLC coupled with a Waters Synapt XS QTOF system. For peptide separation, 5 µg of digested protein was injected onto a reversed-phase charged-sur-face-hybrid C18 column (Waters, PN 186009488) and separated at 45 °C using a water/acetonitrile gradient with 0.1% formic acid.

Reversed-phase separation was followed by TOF MS detection with the following parameters: cone voltage, 40 V; polarity and analyzer mode, positive ion resolution; mass range, 100-2000 m/z; scan rate, 0.5 seconds; collision energy ramp, 20-45 V. Peptides were fragmented for sequencing with collision induced dissociation (CID) using a data independent acquisition method (MSe) acquired by MassLynx software.

The resulting LC-MS/MS data was processed by Protein Metrics Byonic software for peptide mapping, implementing the search parameters for peptide and post-translational modification identification shown in Table 2.

**Table 2.** PMI Byonic Search Parameters.

| Digestion and Instrument Parameters | Value | Details |
| --- | --- | --- |
| Add decoys to search | activated |  |
| Cleavage Sites | E,D | C-terminal for Glu-C digest |
| Digestion Specificity | Fully specific | 2 allowable missed cleavages |
| **Instrument Parameters** |  |  |
| Mass Tolerance | 15 ppm |  |
| Fragment Ion Mass Tolerance | 20 ppm |  |
| Fragmentation Type | QTOF/HCD |  |
| **Modifications** |  |  |
| Deamidation | +0.9840 | Variable, rare 2 @ N,Q |
| Oxidation | +15.9949 | Variable, rare 1 @ M,W |
| Carbamidomethylation | +57.0215 | Variable, common 1 @ C |
| Hexose | +162.0528 | Variable, rare 2 @ K |
| Custom linker-ligand | +1591.83 | Variable, common 2 @ K |
| Cysteinylation | +119.0041 | Variable, common 1 @ C |
| NAcetylCysteine (NAC) | +161.0146 | Variable, common 1 @ C |
| **Spectrum Input Options** |  |  |
| Charge States | 2,3,4,5 |  |
| Precursor Isotope off by x | Too high or low | narrow |
| Maximum precursor mass | 10,000 |  |
| Maximum number of precursors per MS2 | 10 |  |
| Automatic score cut, peptide level | activated |  |
| Protein FDR | 1% | Or 20 reverse sequence count |

Relative abundance of Glu-C peptides was determined manually by MS1 peak area in ion counts. The percentage of amino acid modification was calculated by dividing the ion intensity of the modified peptide by the total ion intensity of all forms of a given peptide (unmodified + modified peptides). Conjugation sites were localized to specific tryptic peptides, however when multiple lysine residues were present on a single tryptic peptide, conjugation sites were ambiguous and not mapped to a specific lysine. To determine the distribution of linker-ligand conjugation across the protein sequence, the ion intensity of a given conjugated peptide was expressed as a percentage of the total intensity of all conjugated peptides.

### In vitro binding assays

#### Determination of MuSK antibody affinities by SPR

Affinities of MuSK antibodies were measured by surface plasmon resonance (Biacore T200 SPR, Cytiva). MuSK antibodies were captured onto a Series S Protein A chip (Cytiva, catalog number 29127556) at a concentration of 0.25 µg/mL for 20 seconds at a rate of 10 µL/min. Association was performed with a series dilution of MuSK Bait or MuSK LYTAC (ranging from 5 nM to 0.125 nM with 2-fold dilutions) for 120 seconds at 30 µL/min, with a subsequent dissociation step for 600 seconds. Regeneration was performed with 10 mM glycine, pH 1.75 or 2.0 for 30 seconds at 30 µL/min for 1 cycle. All reagents were prepared using running buffer (PBS pH 7.4 0.05% Tween-20. Kinetic analysis was performed with Biacore T200 Evaluation Software 3.2.1 using a 1:1 binding model. ***Determination of MuSK antibody affinities by TR-FRET***. Protein and LYTAC were diluted into 1X ultra hi block buffer (Revvity) at various concentrations. Unlabeled MuSK antibodies were then diluted into buffer containing either protein or LYTAC at ∼14 nM and serial diluted into the same buffer. AF647-labeled MuSK antibodies were diluted separately into a buffer containing 0.3 nM Eu anti-human albumin (Revvity, TRF1294C). 15 µl of the unlabeled samples were then added to a 384-well plate (Greiner, 784904) before adding 5 µl of the AF647-labeled MuSK antibodies and allowing them to incubate for 1 hr at 22 °C in the dark. Fluorescence was measured with excitation at 320 nm and emission at 615 nm and 665 nm using a Perkin Elmer EnVision 2105 plate reader.

#### ASGPR-MuSK antibody ternary complex assay

MuSK Bait or MuSK LYTAC were serial diluted 3-fold into binding buffer (20 mM HEPES, 50 mM NaCl, 5 mM CaCl2) and then diluted 1:1 in binding buffer containing

LYTACs for the treatment of myasthenia gravis 10 nM biotinylated-ASGPR (Acro, GS1-H82Q3), 10 nM Tb-streptavidin (Revvity, 610STALB), and 20 nM AF647 labeled anti-MuSK antibody in a 384-well white low volume plate (Greiner, 784904). The mixture was incubated at 22 °C for 1 hr in the dark before measuring fluorescence with excitation at 320 nm and emission at 615 nm and 665 nm using a Perkin Elmer EnVision 2105 plate reader.

### MuSK MG patient samples

#### Patient samples

For MG1-MG10 samples, cryopreserved serum or plasma samples of deidentified patients, with both clinical and laboratory confirmed MuSK autoantibody serostatus, were retrieved from a biorepository established at the Yale University School of Medicine under the approval of Yale University’s Institutional Review Board. Sample MG11 was obtained through Sanguine Biosciences. Clinical and demographic information are provided in Table S2.

#### Patient sera depletion

Control, MuSK Bait and MuSK LYTAC resins were prepared using CNBr-Activated Sepharose 4B (17043001, Cytiva) according to the manufacturer’s instructions. The prepared resins (0.3 mg/mL) were incubated with patient serum samples for 2 hr at 22 °C. Serum samples collected before and after treatment were then assessed in cell binding and AChR clustering assays.

### Cellular assays

#### MuSK-GFP cell binding assay (CBA)

HEK293 cells (ATCC) were transfected in 6-cm cell culture treated dishes (Corning) with DNA encoding MuSK-GFP (Origene Technologies, CW308768) or GFP (Origene, CW308769) using FuGENE 6 (Promega) according to the manufacturer’s instructions. A stable pool of transfected cells was generated through selection with geneticin (Gibco). For cell binding studies, 30,000 cells/well were added to a 96-well conical bottom plate (Thermo Scientific, 14-245-71). Patient samples were diluted 1:20 into FACS buffer (Rockland, MB-089-0500) and serial diluted into the same buffer including equivalent healthy serum. Cells were treated with 1:200 human Fc block (BioLegend, 422302) for 10 minutes before adding diluted samples which were then incubated for 20 minutes at 4 °C. Cells were then washed with FACS buffer and pelleted before addition of goat anti-human IgG AF647 (Invitrogen, A48279). Cells were incubated for 15 minutes at 4 °C, washed with FACS buffer and pelleted before resuspending in FACS buffer and analyzing by flow cytometry (Agilent, NovoCyte Advanteon). Data were analyzed using FlowJo software and the AF647 geometric mean fluorescent intensities were reported.

#### C2C12 AChR clustering assay

C2C12 myoblasts (ATCC) were maintained in growth medium (GM) consisting of high-glucose DMEM without sodium pyruvate (11965092, Thermo Fisher Scientific), supplemented with 10% fetal bovine serum (FBS) (89510-188, Avantor) and 1% penicillin–streptomycin (15140122, Thermo Fisher Scientific). For experiments, myoblasts were seeded at 2,500 cells/well in 96-well cell culture-treated plates, (0720087, Fisher Scientific) or 200,000 cells/well in 6-well, cell culture-treated plates (0720080, Fisher Scientific). Once the cells reached 90% confluence, they were rinsed with PBS, and the GM was replaced with differentiation medium (DM) containing high-glucose DMEM without sodium pyruvate, 2% (v/v) horse serum (26050070, Thermo Fisher Scientific), 1 µM insulin (12585014, Thermo Fisher Scientific), and 1% penicillin–streptomycin. Fresh DM was replenished daily, and the AChR clustering assay was conducted on days 5–6 of differentiation. AChR clustering was induced by adding 10 ng/mL agrin (550-AG-100, R&D Systems) to the DM for 8 (95-well plate) or 16 (6-well plate) hours in the presence or absence of 1 µg/ml MuSK antibody or serum sample, as indicated. After induction, AChR clusters were visualized by staining with 1 µg/mL AF647–labeled α-bungarotoxin (B35450, Invitrogen) for 1 hr at 37 °C. Following staining, cells were washed twice with PBS and fixed with 3% paraformaldehyde (PFA) for 20 minutes at 22 °C. For each treatment, a minimum of four images were captured using an EVOS M7000 (Invitrogen) fluorescence microscope. AChR clusters were quantified using ImageJ software. Background fluorescence was calculated and subtracted from each image, and a consistent threshold was applied across all images to define and count the clusters.

#### Cellular uptake of AF647- or pHrodo green-labeled MuSK antibodies

Hep G2 and Hep G2 *ASGPR*^−/−^ cells were seeded at a density of 50,000 cells per well in a cell-culture treated 96-well plate (Fisherbrand, FB012931) and were cultured for 48 hr at 37 °C prior to experimentation. A mixture of 600 nM AF-647 labeled antibody and 600 nM MuSK LYTAC or MuSK Bait were serial diluted 3-fold into media, as indicated, and incubated for 30 minutes at 22 °C in the dark. Cells were first treated with 50 µL/well human Fc block (BioLegend, 422302) for 20 minutes before adding 50 µL/well of the pre-complexed mixture and incubated for 2 hr at 37 °C. Cells were then harvested with trypLE (Gibco, 12604-021) and transferred into 96-well conical FACS plate (Thermo Scientific, 14-245-71). Cells were washed with FACS buffer and centrifuged at 1200 rpm twice before a final resuspension in FACS buffer and read on the flow cytometer (Agilent, NovoCyte Advanteon). Data analysis was completed using FlowJo software and the mean AF-647 intensity was reported.

Uptake assay using pHrodo-green labeled MuSK antibodies were performed the same as described above, except the cells were incubated with test articles for 24 hr before being harvested.

Data analysis was completed using FlowJo software and the mean pHrodo green intensity was reported. All treatments were run in duplicate or triplicate.

Primary hepatocytes (Sekisui XenoTech, dog D1000. H15B+, cyno PPCH2000+, rat R1000.H15+, human HPCH05+) were seeded at the recommended density for each species in a collagen coated (Gibco, A1048301) 24-well plate (Corning, 07201590) and were cultured for 48 hr at 37 °C prior to experimentation. AF647-labeled MuSK antibody uptake procedures were performed as described above, apart from using canine Fc block (Invitrogen, 14-9162-42) for dog hepatocytes and mouse Fc block (Biolegend, 101320) for rat hepatocytes instead.

#### MuSK antibody degradation

##### Quenched TAMRA probe degradation assay

To generate the quenched TAMRA probe, a goat anti-human IgG F(ab′)2 fragment (Jackson ImmunoResearch 109-006-170) was labeled with QSY 7 succinimidyl ester (ThermoFisher, Q10193) and TAMRA succinimidyl ester (ThermoFisher, C1171) and purified as previously described (37). Hep G2 cells were seeded at a density of 200,000 cells per well in a cell-culture treated 24-well plate (Corning, 3524) and were cultured for 48 hr at 37 °C prior to experimentation. A mixture of 50 nM each quenched TAMRA probe, MuSK Bait or LYTAC (as indicated), and MuSK antibodies were pre-complexed for 30 minutes at 22 °C. Media from cells was then aspirated before adding 300 µl/well of the pre-complexed mixture to the cells. Cells were imaged using an IncuCyte S3 microscope (Sartorius) at 37 °C where three pictures per well were taken every hour for three days. Images were analyzed using the IncuCyte 2020A software package.

##### LLDR degradation assay

Hep G2 and Hep G2 *ASGPR*^−/−^ cells were seeded at a density of 50,000 cells per well in a cell-culture treated 96-well plate (Fisherbrand, FB012931) and were cultured for 48 hr at 37 °C prior to experimentation. A mixture of 600 nm LLDR-antibody and 600 nM MuSK LYTAC or MuSK Bait were serial diluted 3-fold into media, as indicated, and incubated for 30 minutes at 22 °C in the dark. Cells were first treated with 50 µL/well human Fc block (BioLegend, 422302) for 20 minutes before adding 50 µL/well of the pre-complexed mixture and incubated for 2 hr at 37 °C. Cells were then trypsinized with 50 µL/well trypLE (Gibco, 12604-021) for 8-10 minutes and transferred into 96-well FACS plates (Thermo Scientific, 14-245-71). Cells were washed with FACS buffer (Rockland, MB-089-0500) and centrifuged twice before final resuspension in FACS buffer and read on the flow cytometer (Agilent, NovoCyte Advanteon). Data analysis was completed using FlowJo software and the mean LLDR intensity was reported.

### Animal handling and general procedures

All animal studies were performed in an AAALAC approved facility under Institutional Animal Care and Use Committee guidelines. Rat PK studies were conducted male Sprague-Dawley rats at Charles River Labs, South San Francisco. Age-matched female C57Bl/6 mice were procured from either Charles River Laboratories or The Jackson Laboratory. NOD SCID mice (NOD.Cg-*Prkdc*^*scid*^/J) were obtained from The Jackson Laboratory. Mice were co-housed in ventilated, disposable cages (Innovive, San Diego, CA). Food and water were provided ad libitum.

During PK and pharmacodynamic studies, MuSK antibody was prepared fresh from stock and dosed IV. MuSK-LYTAC and MuSK Bait was dosed either IV or SC, as indicated, and efgartigimod was dosed IP. Serum was isolated following tail vein blood collection. Harvest of liver, kidney and spleen were performed immediately following perfusion and tissues were flash frozen for downstream processing.

For passive immunization studies, MuSK antibody was administered IP. MuSK-LYTAC was dosed subcutaneously and efgartigimod was dosed IP. Body weight was monitored three to five times per week initially, then daily, once weight loss was observed. Treatment groups were terminated once weight loss exceeded 20 percent. Forelimb muscle weakness was determined using a BIOSEB (North Pinellas Park, Florida) GT4 grip strength meter. Five replicate grip strength measurements were performed on each mouse and averaged. Statistical analysis of studies was performed using GraphPad Prism software. PK parameters were determined using WinNonlin.

### Tissue processing

Livers and kidneys were sliced into thin sections and transferred to 5 mL Eppendorf tubes prefilled with stabilized ceramic beads (NA-RED5E100, Stellar Scientific) and 2 mL of lysis buffer. The lysis buffer was prepared using Tissue Extraction Reagent 1 (FNN0071, Thermo Fisher Scientific) supplemented with Halt Protease and Phosphatase Inhibitor Cocktail 100x (78446, Thermo Fisher Scientific), 0.05 mM PMSF Protease Inhibitor (36978, Life Technologies), and 25 U of Pierce Universal Nuclease (88701, Thermo Fisher Scientific). Tissues were homogenized using a Bullet Blender (NA-BT5E, Stellar Scientific) at speed 8 for 3 minutes.

For spleen homogenization, spleens were placed on a cell strainer in a small Petri dish containing 1 mL of lysis buffer. A syringe with a flat plunger was used to press the spleens through the strainer into the dish, followed by rinsing the strainer with an additional 1 mL of lysis buffer. The homogenized spleen mixture (2 mL of lysis buffer) was transferred to 5 mL Eppendorf tubes prefilled with ceramic beads and homogenized using the Bullet Blender at speed 8 for 3 minutes.

All tissue lysates were centrifuged at 10,000 rpm for 5 minutes at 4 °C. The resulting supernatants were further centrifuged at maximum speed for 20 minutes at 4 °C, and the final supernatants were collected. Total protein concentrations were measured using the BCA Protein Assay Kit (23227, Life Technologies) according to the manufacturer’s instructions.

### ELISA detection methods

For all the methods described below, plates were coated for 16 hr at 4 °C with the indicated antibodies. Subsequent steps were conducted at 22 °C. Samples were developed with tetramethylbenzidine substrate (Sigma Aldrich, T0440-1L), quenched with ELISA stop solution (Invitrogen, SS04) and absorbance at 450 nm was measured using a Perkin Elmer EnVision 2105 plate reader. Analyte levels were determined by interpolation from standard curves that were fit to a four-parameter fit model in GraphPad Prism.

### ELISA detection of biotinylated MuSK antibodies

MaxiSorp 96-well plates (Thermo Scientific, 442404) coated overnight at 4 °C with 2 µg/mL goat anti-human Fc (Thermo, 31125) were blocked with PBS with 0.05% Tween-20 (PBS-T) and 1% BSA for 1 hr. Serum samples diluted in PBS-T 1% BSA 1 µg/mL MuSK-HSA were serial diluted into the same buffer including equivalent naïve serum and pipetted into the wells and incubated for 1 hr. After washing in PBS-T, a 1:2000 dilution of streptavidin-HRP (Pierce, 21130) was added into the wells for 1 hr before washing and developing the plates.

### ELISA detection of MuSK-HSA in mouse serum

MaxiSorp 96-well plates coated overnight at 4 °C with 2 µg/mL mouse anti-HSA (Invitrogen, MA120127) were blocked with PBS-T 1% BSA for 1 hr. Serum samples diluted in PBS-T 1% BSA were serial diluted into the same buffer including equivalent naïve serum and pipetted into the wells and incubated for 1 hr. After washing in PBS-T, 2 µg/mL goat anti-HSA (Southern Biotech, 2080-01) was added into the wells for 1 hr. After washing, a 1:4000 dilution of bovine anti-goat IgG HRP (Jackson ImmunoResearch, 805-035-180) was added into the wells for 1 hr before washing and developing the plates.

### ELISA detection of MuSK-HSA in rat serum

MaxiSorp 96-well plates coated overnight at 4 °C with 2 µg/mL MuSK antibody 11-3F6 were blocked with PBS-T 1% BSA for 1 hr. Serum samples diluted in PBS-T 1% BSA were serial diluted into the same buffer including equivalent naïve rat serum and added to the plate and incubated for 1 hr. After washing in PBS-T, 1 µg/mL biotinylated MuSK1A was added into the wells for 1 hr. After washing, a 1:2000 dilution of streptavidin-HRP (Pierce, 21130) was added into the wells for 1 hr before washing and developing the plates.

### ELISA detection of monovalent MuSK antibodies

MaxiSorp 96-well plates coated overnight at 4 °C with 10 µg/mL hemocyanin from keyhole limpet (KLH) (Sigma Aldrich, H8283) were blocked with PBS-T 1% BSA for 1 hr. Serum samples diluted in PBS-T 1% BSA were serial diluted into the same buffer including equivalent naïve serum, pipetted into the wells, and incubated for 1 hr. 5 µg/ mL MuSK HSA was added into the dilution buffer in samples that had been treated with LYTAC or protein. After washing in PBS-T, a 1:2000 dilution of goat anti-human IgG HRP (Sigma Aldrich, SAB3701283) was added into the wells for 1 hr before washing and developing the plates.

### ELISA detection of bivalent MuSK antibodies

MaxiSorp 96-well plates coated overnight at 4 °C with 2 µg/mL goat anti-human Fc (Thermo, 31125) were blocked with PBS-T 1% BSA for 1 hr. Serum samples diluted in PBS-T 1% BSA were serial diluted into the same buffer including equivalent naïve serum, pipetted into the wells, and incubated for 1 hr before washing and developing the plates.

### ELISA detection of total mouse IgG

Serum samples were diluted 1:100 and were assayed and analyzed based on manufacturer’s instructions from the total mouse IgG ELISA detection kit (Invitrogen, 88-50400-88).

### ELISA detection of conjugated MuSK-HSA

MaxiSorp 96-well plates coated overnight at 4 °C with 2 µg/mL anti-MuSK antibody 11-3F6 (in-house) were blocked with 20 mM Tris 150 mM NaCl 0.05% Tween-20 5 mM CaCl2 (TBS-T-Ca) 1% BSA for 1 hr. Serum samples diluted in TBS-T-Ca 1% BSA were serial diluted into the same buffer including equivalent naïve serum, pipetted into the wells, and incubated for 1 hr. After washing in TBS-T-Ca, 2 µg/mL biotinylated ASGPR (Acro, GS1-H82Q3) was added into the wells for 1 hr. After washing in TBS-T-Ca, 1:2000 dilution of streptavidin-HRP (Pierce, 21130) was added into the wells for 1 hr before washing and developing the plates.

### List of Supplementary Materials

Supplemental Materials and Methods Fig S1 to S14

Table S1 to S2

## Data Availability

All data produced in the present study are available upon reasonable request to the authors

## Author contributions

Conceptualization: NAL, RK, TTT, STS, DAL, GM, KCO, SMM, JSI

Methodology: NAL, CLC, CP, RK, SMT, DL, TC, RML, TTT, GM, EDT, DAL, KCO, SMM, JSI

Investigation: NAL, CLC, CP, RK, SMT, DL, TC, RML, TTT, CC, EDT

Visualization: NAL, RK, SMT, SMM, JSI

Supervision: NAL, TTT, RJN, STS, EDT, DAL, KCO, SMM, JSI

Writing – original draft: NAL, CLC, CP, RK, SMT, DL, TC, RML, EDT, KCO, SMM, JSI

Writing – review & editing: NAL, CLC, CP, RK, SMT, DL, TC, RML, TTT, CC, GM, RJN, STS, EDT, DAL, KCO, SMM, JSI

