## Supplemental Materials for "Selective degradation of pathogenic autoantibodies by lysosomal targeting chimeras for the treatment of myasthenia gravis"

**Supplementary Materials**

**Materials and Methods**

**Chemical synthesis**

Except where noted, chemicals that are commercially available were purchased from Sigma-Aldrich, CombiBlocks, TCI, BroadPharma or Enamine and were used without further purification. Liquid chromatography–mass spectrometry (LCMS) was carried out on a ThermoScientific LTQXL instrument equipped with a Gemini NX-C18 column (50 x 2 mm, 3µm) and high-resolution MS on a Waters Synapt XS instrument. Flash column chromatography was performed on a Teledyne ISCO Combiflash Rf Lumen installed with disposable normal phase RediSep Rf columns (SiliCycle). Preparative HPLC purification was performed on an ACCQPrep HP150 preparative HPLC system equipped with a Gemini NX-C18 column (250 mm × 50 mm and 10 μm particle size, Phenomenex) using gradients as indicated. Compound characterization using NMR was performed on a Bruker 400 mHz AvanceCore spectrometer. Signal patterns are described as singlet (s), doublet (d), triplet (t), quartet (q), multiplet (m), broad singlet (bs), or a combination of the listed splitting patterns. The coupling constants (*J*) are measured in hertz (Hz).

**Synthesis of activated ASGPR linker ligand 1**
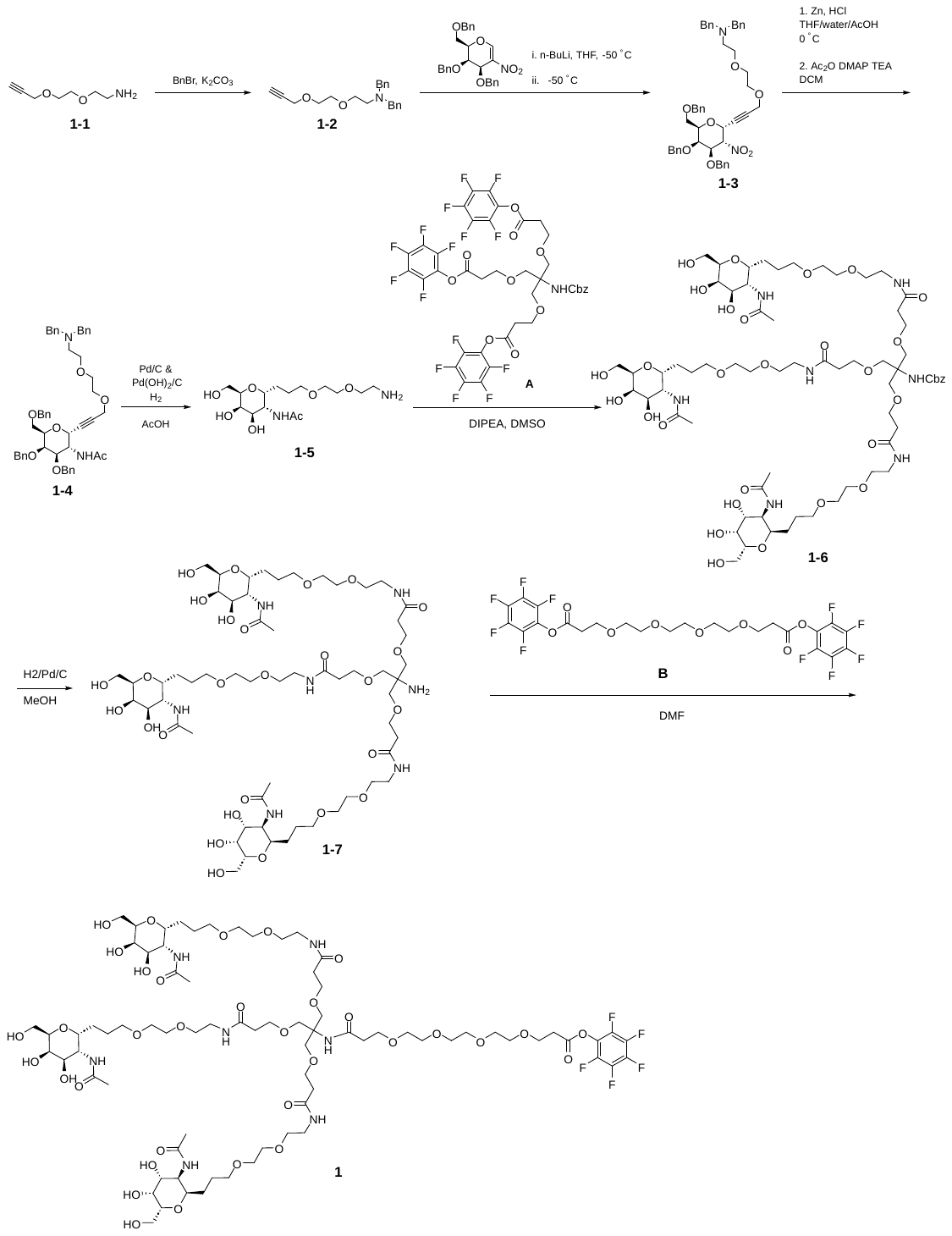

*N,N-Dibenzyl-2-(2-(prop-2-yn-1-yloxy)ethoxy)ethan-1-amine* (**1-2**)

A mixture of 2-(2-(prop-2-yn-1-yloxy)ethoxy)ethan-1-amine (**1-1**, 1.00 eq, 9.75 g, 68.1 mmol), anhydrous potassium carbonate (2.32 eq, 21.82 g, 158 mmol) and acetonitrile (306.46 mL) was treated with benzyl bromide (2.10 eq, 17 mL, 143 mmol), then heated to 50 °C for 2 hr. The reaction was filtered through celite and the filtrate was concentrated under vacuum. The residue was purified by column chromatography (5-100% EtOAc in hexanes) to give **1-2** as clear oil. Yield: 18.4 g, 83%; LCMS m/z 324.22 [M+H]^+^, ^1^H NMR (400 MHz, CDCl3) δ 7.37 (d, J = 7.0 Hz, 4H), 7.29 (dd, J = 8.3, 6.6 Hz, 4H), 7.21 (t, J = 7.0 Hz, 2H), 4.17 (d, J = 2.4 Hz, 2H), 3.66 – 3.62 (m, 6H), 3.59 – 3.53 (m, 4H), 2.70 (t, J = 6.3 Hz, 2H), 2.38 (t, J = 2.4 Hz, 1H). HRMS (m/z): [M+H]^+^ calculated for C21H25NO2, 324.1958; found, 324.1944.

*N,N-dibenzyl-2-(2-((3-((2R,3S,4R,5R,6R)-4,5-bis(benzyloxy)-6-((benzyloxy)methyl)-3-nitrotetrahydro-2H-pyran-2-yl)prop-2-yn-1-yl)oxy)ethoxy)ethan-1-amine* (**1-3**)

In an oven dried flask, a solution of N,N-dibenzyl-2-(2-(prop-2-yn-1-yloxy)ethoxy)ethan-1-amine (**1-2**, 1.43 eq, 3.70 g, 11.4 mmol) in anhydrous THF (31.8 mL) under nitrogen via balloon was cooled to -50 °C (dry-ice bath - 1:1 MeOH/water) then treated with the slow addition of a solution of 2.5M butyl lithium in hexane (1.20 eq, 3.8 mL, 9.62 mmol) and the reaction was stirred at -50 °C for 60 minutes. Next, a solution of (2R,3R,4R)-3,4-dibenzyloxy-2-(benzyloxymethyl)-5-nitro-3,4-dihydro-2H-pyran (1.00 eq, 3.70 g, 8.02 mmol) in dry THF (12.7 mL) was added dropwise over 5 minutes while maintaining -50 °C externally. After 60 minutes, the reaction was quenched with the addition of 7.1M aq. ammonium chloride (32.8 eq, 37 mL, 263 mmol) until a reaction pH of 9-10 was reached, then the cold bath was removed, and the slurry was warmed to room temperature. The desired product was extracted with EtOAc (100 mL, 50 mL) and the combined organic layers were dried over Na_2_SO_4_, filtered and concentrated under reduced pressure to give a crude oil. The oil was adsorbed to silica gel then purified by silica gel chromatography (10% then 20% 2-MeTHF in hexanes) to give **1-3** as the desired product. Yield: 2.19 g, 35 %; LCMS m/z 785.34 [M+H]^+^, ^1^H NMR (400 MHz, CDCl3) δ 7.39 – 7.13 (m, 25H), 5.28 (dt, J = 6.0, 1.8 Hz, 1H), 5.12 (dd, J = 10.6, 5.9 Hz, 1H), 4.82 (d, J = 11.1 Hz, 1H), 4.68 (s, 2H), 4.52 – 4.31 (m, 4H), 4.21 (dd, J = 4.9, 1.8 Hz, 2H), 4.12 (t, J = 6.6 Hz, 1H), 4.02 (d, J = 2.9 Hz, 1H), 3.63 (s, 4H), 3.62 – 3.47 (m, 8H), 2.69 (t, J = 6.3 Hz, 2H). ^13^C NMR (101 MHz, CDCl3) δ 139.76, 137.86, 137.60, 137.09, 128.75, 128.52, 128.48, 128.36, 128.17, 128.11, 127.97, 127.93, 127.88, 126.83, 87.17, 83.71, 77.56, 77.22, 76.21, 75.22, 73.60, 73.04, 73.00, 72.80, 70.05, 70.01, 69.28, 68.05, 66.57, 58.94, 58.32, 52.68, 30.32. HRMS (m/z): [M+H]^+^ calculated for C48H52N2O8, 785.3797; found, 785.3780.

*N-((2R,3S,4R,5R,6R)-4,5-bis(benzyloxy)-6-((benzyloxy)methyl)-2-(3-(2-(2-(dibenzylamino)ethoxy)ethoxy)prop-1-yn-1-yl)tetrahydro-2H-pyran-3-yl)acetamide* (**1-4**)

A solution of N,N-dibenzyl-2-(2-((3-((2R,3S,4R,5R,6R)-4,5-bis(benzyloxy)-6-((benzyloxy)methyl)-3-nitrotetrahydro-2H-pyran-2-yl)prop-2-yn-1-yl)oxy)ethoxy)ethan-1-amine (**1-3**, 1.00 eq, 2.40 g, 3.06 mmol) in THF (182 mL), water (77.6 mL) and acetic acid (46.1 mL) was cooled over ice then treated with zinc dust (18.8 eq, 3.76 g, 57.5 mmol) followed by 12M HCl (44.3 eq, 11 mL, 135 mmol). After 90 minutes, the reaction was filtered then re-cooled in an ice bath before it was treated with 5M aq. sodium hydroxide (350 eq, 214 mL, 1070 mmol) at such a rate as to keep the internal temperature below 24 °C. Next, the layers were partitioned then the aqueous layer was extracted with DCM (80 mL). The aqueous layer was extracted with DCM (50 mL) then the combined organic layers were washed with brine then dried over Na_2_SO_4_, filtered, concentrated under reduced pressure and left under high vacuum.

The crude amine (2.3 g) was then dissolved in DCM (20 mL) before adding triethylamine (6.00 eq, 2.6 mL, 18.3 mmol), 4-(dimethylamino)pyridine (0.050 eq, 18.7 mg, 0.153 mmol) then acetic anhydride (9.80 eq, 2.8 mL, 30.0 mmol). After several hr, the reaction was quenched with water (20 mL) for several minutes. The organic layer was collected, and the aqueous layer was extracted with DCM (2x10 mL). The combined organic layers were dried over Na_2_SO_4_, filtered and concentrated onto silica gel for purification by silica gel chromatography (0-15% 2-MeTHF in DCM) to give **1-4** as the desired product. Yield: 1.86 g, 76%. LCMS m/z 797.5 [M+H]^+^. ^1^H NMR (400 MHz, CDCl3) δ 7.37 – 7.22 (m, 26H), 5.09 (d, J = 7.5 Hz, 1H), 5.07 – 5.01 (m, 1H), 4.88 (d, J = 11.9 Hz, 1H), 4.71 (d, J = 12.2 Hz, 1H), 4.60 – 4.52 (m, 2H), 4.46 (q, 2H), 4.37 (d, J = 12.2 Hz, 1H), 4.15 (s, 2H), 4.06 – 3.99 (m, 2H), 3.62 – 3.51 (m, 12H), 2.68 (t, J = 6.5 Hz, 2H), 1.83 (d, J = 2.5 Hz, 3H). HRMS (m/z): [M+H]^+^ calculated for C50H56N2O7, 797.4161; found, 797.4147.

*N-((2R,3R,4R,5R,6R)-2-(3-(2-(2-aminoethoxy)ethoxy)propyl)-4,5-dihydroxy-6-(hydroxymethyl)tetrahydro-2H-pyran-3-yl)acetamide* (**1-5**)

A mixture of N-((2R,3S,4R,5R,6R)-4,5-bis(benzyloxy)-6-((benzyloxy)methyl)-2-(3-(2-(2-(dibenzylamino)ethoxy)ethoxy)prop-1-yn-1-yl)tetrahydro-2H-pyran-3-yl)acetamide (**1-4**, 1.00 eq, 2.02 g, 2.53 mmol), and 10% Pd/C w/w (dry basis) (0.040 eq, 0.22 g, 0.101 mmol) in acetic acid (86 mL) under nitrogen was evacuated then back-filled with hydrogen gas via balloon - a process that was repeated 3 times before leaving under an atmosphere of hydrogen. After 1 hr, 10% Pd/C w/w (dry basis) (0.200 eq, 1.08 g, 0.507 mmol) and 20% w/w (dry basis) palladium hydroxide (0.300 eq, 1.07 g, 0.760 mmol) were added and the reaction was again placed under hydrogen. After 3 more hr, the reaction was filtered over a pad of celite then the filter cake was rinsed while stirring with methanol (100 mL). Solvents were removed under reduced pressure and the residue was concentrated from toluene then left under high vacuum. The residue was purified by HPLC (eluting from a C18 column with 5-20% ACN in water w/ 20mM NH4OH) and fractions lyophilized to give **1-5** as a white solid. Yield: 562 mg, 63%. LCMS m/z 351.4 [M+H]^+^; ^1^H NMR (400 MHz, DMSO-*d*6 + D2O) δ 3.99 (dd, J = 9.2, 4.9 Hz, 1H), 3.83 (dt, J = 9.9, 4.4 Hz, 1H), 3.72 (d, J = 2.9 Hz, 1H), 3.61 – 3.45 (m, 8H), 3.37 (q, J = 6.1 Hz, 4H), 2.63 (t, J = 5.8 Hz, 2H), 1.83 (s, 3H), 1.58 (tq, J = 13.4, 4.3 Hz, 2H), 1.34 (dddt, J = 38.6, 13.7, 10.1, 5.5 Hz, 2H). ^13^C NMR (101 MHz, DMSO-d6) δ 170.04, 73.57, 73.02, 71.33, 70.62, 70.02, 69.86, 68.18, 67.81, 60.28, 50.54, 41.36, 26.11, 23.02, 22.88. HRMS (m/z): [M+H]^+^ calculated for C15H30N2O7, 351.2126; found, 351.2114.

*Benzyl (1,31-bis((2R,3R,4R,5R,6R)-3-acetamido-4,5-dihydroxy-6-(hydroxymethyl)tetrahydro-2H-pyran-2-yl)-16-(15-((2R,3R,4R,5R,6R)-3-acetamido-4,5-dihydroxy-6-(hydroxymethyl)tetrahydro-2H-pyran-2-yl)-5-oxo-2,9,12-trioxa-6-azapentadecyl)-11,21-dioxo-4,7,14,18,25,28-hexaoxa-10,22-diazahentriacontan-16-yl)carbamate* (**1-6**)

A mixture of N-((2R,3R,4R,5R,6R)-2-(3-(2-(2-aminoethoxy)ethoxy)propyl)-4,5-dihydroxy-6-(hydroxymethyl)tetrahydro-2H-pyran-3-yl)acetamide (**1-5**, 3.24 eq, 1.72 g, 4.91 mmol) and bis(perfluorophenyl) 3,3'-((2-(((benzyloxy)carbonyl)amino)-2-((3-oxo-3-(perfluorophenoxy)propoxy)methyl)propane-1,3-diyl)bis(oxy))dipropionate (**A**, 1.00 eq, 1.47 g, 1.52 mmol) was treated with diisopropylethylamine (3.00 eq, 862 mL, 4.95 mmol) then dissolved in DMSO (6.89 mL). After stirring for 90 minutes, the reaction was directly purified by HPLC (5%-50% acetonitrile in water w/0.1% FA) to give **1-6** as a white solid. Yield: 2.21 g, 91%. LCMS m/z 1468.6 [M+H]^+^; ^1^H NMR (400 MHz, CD_3_OD) δ 7.41 – 7.26 (m, 5H), 5.04 (s, 2H), 4.25 (dd, *J* = 9.6, 5.1 Hz, 3H), 4.10 (dt, *J* = 10.9, 4.4 Hz, 3H), 3.90 (t, *J* = 2.8 Hz, 3H), 3.81 (dd, *J* = 12.6, 8.3 Hz, 3H), 3.75 – 3.63 (m, 21H), 3.61 – 3.56 (m, 12H), 3.55 – 3.48 (m, 12H), 3.38 – 3.33 (m, 6H), 2.43 (t, *J* = 6.1 Hz, 6H), 1.98 (s, 9H), 1.81 – 1.66 (m, 6H), 1.62 – 1.39 (m, 6H).

*1,31-bis((2R,3R,4R,5R,6R)-3-acetamido-4,5-dihydroxy-6-(hydroxymethyl)tetrahydro-2H-pyran-2-yl)-16-(15-((2R,3R,4R,5R,6R)-3-acetamido-4,5-dihydroxy-6-(hydroxymethyl)tetrahydro-2H-pyran-2-yl)-5-oxo-2,9,12-trioxa-6-azapentadecyl)-11,21-dioxo-4,7,14,18,25,28-hexaoxa-10,22-diazahentriacontan-16-amine*  (**1-7**)

A solution of **1-6** (1.00 eq, 400 mg, 0.272 mmol) in methanol (20 mL) was purged with nitrogen then treated with 10% Pd/C (0.400 eq, 232 mg, 0.109 mmol) before being evacuated then back-filled with hydrogen via balloon. After 2 hr, the reaction was treated with 500 mg celite then filtered over a pad of celite. The filter cake was rinsed with methanol and the filtrate was concentrated under reduced pressure. The residue was dissolved in water with minimal DMSO and then purified by reversed-phase HPLC (5-50% acetonitrile in water w/ 0.2 mM NH_4_OH) to give **1-7**. Yield: 315 mg, 86.7%. LCMS m/z 1334.6 [M+H]^+^; ^1^H NMR (400 MHz, CD_3_OD) δ 4.27 (dd, *J* = 9.9, 4.8 Hz, 3H), 4.15 – 4.08 (m, 3H), 3.91 (t, *J* = 2.8 Hz, 3H), 3.83 (dd, *J* = 12.4, 8.6 Hz, 3H), 3.78 – 3.68 (m, 15H), 3.65 – 3.51 (m, 24H), 3.40 (t, *J* = 5.5 Hz, 6H), 3.38 – 3.35 (m, 6H), 2.47 (t, *J* = 6.1 Hz, 6H), 2.00 (s, 9H), 1.84 – 1.70 (m, 6H), 1.65 – 1.43 (m, 6H).

*Perfluorophenyl 33-((2R,3R,4R,5R,6R)-3-acetamido-4,5-dihydroxy-6-(hydroxymethyl)tetrahydro-2H-pyran-2-yl)-18,18-bis(15-((2R,3R,4R,5R,6R)-3-acetamido-4,5-dihydroxy-6-(hydroxymethyl)tetrahydro-2H-pyran-2-yl)-5-oxo-2,9,12-trioxa-6-azapentadecyl)-16,23-dioxo-4,7,10,13,20,27,30-heptaoxa-17,24-diazatritriacontanoate* (**1**)

A solution of **1-7** (1.00 eq, 1.10 g, 0.824 mmol) in DMF (13.7 mL) was added dropwise to a solution of bis(perfluorophenyl) 4,7,10,13-tetraoxahexadecanedioate (**B**, 3.00 eq, 1.55g, 2.47 mmol) in DMF (5.6 mL) via syringe over 16 minutes. Next, DMF (2 mL) was added to the amine flask for rinsing then this solution was added to the reaction dropwise via syringe. The reaction was stirred at room temperature overnight. After 26 hr, the reaction was purified directly by reversed-phase HPLC (5-40% acetonitrile in water w/0.1% TFA) to give **1** as a solid. Yield: 1050 mg, 71%. LCMS m/z 1776.5 [M+H]^+^; ^1^H NMR (400 MHz, D_2_O) δ 4.13 (dd, *J* = 10.9, 5.9 Hz, 3H), 4.06 – 3.97 (m, 3H), 3.91 – 3.84 (m, 5H), 3.81 (dd, *J* = 10.9, 3.3 Hz, 3H), 3.71 – 3.62 (m, 21H), 3.63 – 3.56 (m, 26H), 3.54 (t, *J* = 5.5 Hz, 6H), 3.49 (t, *J* = 6.4 Hz, 6H), 3.32 (t, *J* = 5.5 Hz, 6H), 3.00 (t, *J* = 5.9 Hz, 2H), 2.42 (t, *J* = 6.0 Hz, 8H), 1.94 (s, 9H), 1.76 – 1.58 (m, 6H), 1.54 – 1.35 (m, 6H); ^13^C NMR (101 MHz, DMSO) δ 170.34, 170.28, 169.41, 167.90, 141.95 - 136.10 (m, 3C), 124.40 (m, 1C), 73.33, 70.74, 70.26, 69.81, 69.76, 69.71, 69.69, 69.63, 69.59, 69.48, 69.37, 69.16, 68.26, 67.97, 67.45, 67.33, 66.86, 65.59, 59.91, 59.65, 50.29, 38.53, 36.60, 35.88, 33.98, 25.70, 22.64, 22.56; ^19^F NMR (376 MHz, DMSO) δ -153.28 (d, *J* = 22.8 Hz, 2F), -158.05 (t, *J* = 23.2 Hz, 1F), -162.65 (dd, *J* = 23.3, 19.1 Hz, 2F); HRMS (*m/z*): [M+H]^+^ calculated for C76H127F5N7O34, 1776.8339; found, 1776.8322.

**Synthesis of enantiomeric ASGPR linker ligand control (2)**

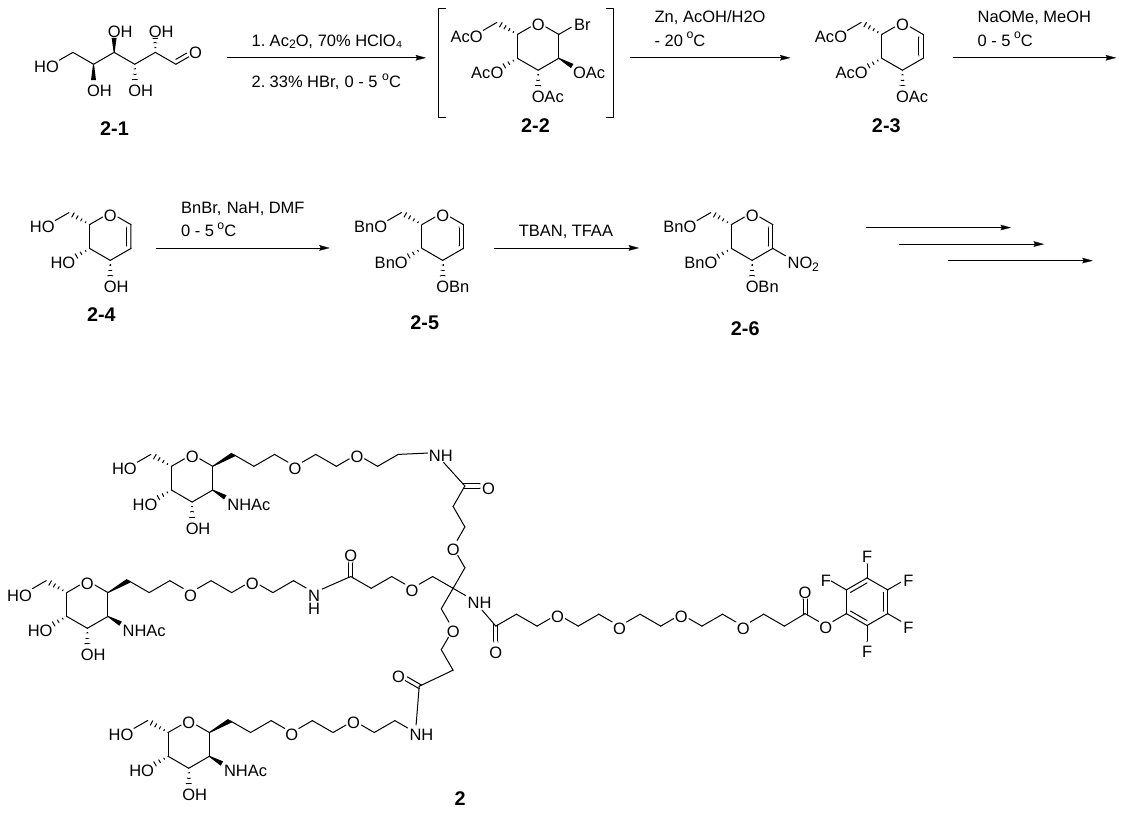

*Synthesis of (2S,3S,4S)-2-(acetoxymethyl)-3,4-dihydro-2H-pyran-3,4-diyl diacetate* (**2-3**)

To a solution of acetic anhydride (6.3 eq, 65 mL, 688 mmol) was slowly added 70% perchloric acid (0.2 eq, 1.9 mL, 22 mmol), while keeping solution at ambient temperature. While stirring, (2S,3R,4R,5S)-2,3,4,5,6-pentahydroxyhexanal (**2-1**) (1.0 eq, 19.6 g, 108 mmol) was added portion-wise as a solid over approximately 30 minutes. The reaction stirred at ambient temperature for approximately 2 hr, then the reaction mixture was cooled to 0-5 ^o^C, and 33% w/w hydrogen bromide in acetic acid (3.5 eq, 66.0 mL, 377 mmol) was added dropwise over 30 minutes. The reaction continued to stir while warming to ambient temperature. After 2 hr, the reaction solution was diluted with dichloromethane, transferred to a separatory funnel, and washed with 2 x H_2_O, 2 x 5% w/w aqueous sodium bicarbonate, and 1 x brine. The dichloromethane layer was dried over sodium sulfate and solvent evaporated to afford crude (2S,3R,4R,5S)-2-(acetoxymethyl)-6-bromotetrahydro-2H-pyran-3,4,5-triyl triacetate (**2-2**) as a dark brown color syrup. The crude **2-2** was dissolved in 50% v/v acetic acid/H_2_O (150 mL), stirred while cooling to approximately -20 ^o^C, and zinc dust (35.8 g, 548 mmol, 5.0 eq.) was added as a solid portion-wise over approximately 30 minutes. The reaction was stirred while warming to ambient temperature. After approximately 30 minutes, solids were removed by filtration over diatomaceous earth, followed by washing with dichloromethane. The filtrate was transferred to a separatory funnel, layers were separated, lower dichloromethane layer was washed with 2 x H_2_O and 1 x brine, dried over sodium sulfate, and solvent evaporated to crude syrup. The product was isolated by silica chromatography, eluting with 0-10% ethyl acetate/dichloromethane. Fractions containing product were combined, solvent evaporated, and dried under high-vacuum at ambient temperature to **2-3** as a clear syrup. Yield: 15.1 g, 51%; LCMS m/z 295.03 [M+Na]^+^; ^1^H NMR (400 MHz, CDCl_3_) δ 7.25 (d, *J* = 6.3 Hz, 1H), 6.39 – 6.14 (m, 2H), 5.58 – 5.43 (m, 1H), 5.18 – 4.87 (m, 3H), 2.92 (s, 3H), 2.88 (s, 3H), 2.82 (s, 3H).

*(2S,3S,4S)-2-(hydroxymethyl)-3,4-dihydro-2H-pyran-3,4-diol* (**2-4**)

A solution of (2S,3S,4S)-2-(acetoxymethyl)-3,4-dihydro-2H-pyran-3,4-diyl diacetate (**2-3**, 1.0eq, 14.9 g, 54.7 mmol) in methanol (150 mL) was cooled to 0-5 ^o^C while stirring, followed by slow addition of 25% w/w sodium methoxide in methanol (0.3 eq, 3.8 mL, 16.6 mmol). The reaction mixture was stirred while warming to ambient temperature. After approximately 15 minutes, the reaction mixture was evaporated onto silica and the product isolated by silica chromatography, eluting with 70-100% ethyl acetate/dichloromethane, followed by product elution with 0-20% methanol/ethyl acetate. Fractions containing product were combined, the solvent evaporated, and dried under high-vacuum at ambient temperature to afford **2-4** as a white solid. Yield: 6.9 g, 86%; LCMS m/z 169.0 [M+Na]^+^; ^1^H NMR (400 MHz, CD_3_OD) δ 6.34 (dd, *J* = 6.3, 1.9 Hz, 1H), 4.62 (dt, *J* = 6.3, 2.0 Hz, 1H), 4.36 – 4.30 (m, 1H), 3.94 – 3.86 (m, 2H), 3.86 – 3.70 (m, 2H). [α]_D_^20^=15.42^o^ (c 1.200, MeOH).

*(2S,3S,4S)-3,4-bis(benzyloxy)-2-((benzyloxy)methyl)-3,4-dihydro-2H-pyran* (**2-5**)

A solution of (2S,3S,4S)-2-(hydroxymethyl)-3,4-dihydro-2H-pyran-3,4-diol (**2-4**) (1.0 eq, 6.61 g 45.2 mmol) in DMF (60 mL) was stirred while cooling to 0-5 ^o^C, followed by portion-wise addition of sodium hydride (60% dispersion in mineral oil) (4.0 eq, 7.28 g, 182 mmol). The slurry mixture continued to stir at 0-5 ^o^C for approximately 40 minutes, then benzyl bromide (4.0 eq, 22.0 mL, 185 mmol) was added dropwise over approximately 25 minutes. The reaction slurry continued to stir while warming to ambient temperature. After approximately 90 minutes, the reaction slurry was again cooled to 0-5 ^o^C and quenched with dropwise addition of methanol (7 mL) and the resulting solution stirred for 15 minutes. The solution was then diluted with ethyl acetate, transferred to separatory funnel, and washed twice with cold water and once with brine, dried over sodium sulfate, and solvent evaporated to a crude syrup. The product was purified by silica chromatography, eluting with 0-50% ethyl acetate/hexanes. Fractions containing product were combined, the solvent evaporated and dried under high-vacuum at ambient temperature to afford **2-5** as a white solid. Yield: 15.8 g, 84%; LCMS m/z 439.2 [M+Na]^+^; ^1^H NMR (400 MHz, DMSO) δ 7.39 – 7.22 (m, 15H), 6.34 (dd, *J* = 6.2, 1.7 Hz, 1H), 4.84 – 4.74 (m, 2H), 4.62 (s, 2H), 4.56 (d, *J* = 11.5 Hz, 1H), 4.52 – 4.41 (m, 2H), 4.29 – 4.16 (m, 2H), 4.03 – 3.96 (m, 1H), 3.66 (qd, *J* = 10.3, 6.0 Hz, 2H).

*(2S,3S,4S)-3,4-bis(benzyloxy)-2-((benzyloxy)methyl)-5-nitro-3,4-dihydro-2H-pyran* (**2-6**)

A solution of (2S,3S,4S)-3,4-bis(benzyloxy)-2-((benzyloxy)methyl)-3,4-dihydro-2H-pyran (**2-5**) (1.0 eq, 9.70 g, 23.3 mmol) in dichloromethane (200 mL) was added tetrabutylammonium nitrate (1.2eq, 8.53 g, 28.0 mmol). The solution was stirred under nitrogen atmosphere while cooling to 0-5 °C, then trifluoroacetic anhydride (1.2 eq, 3.94 mL, 28.3 mmol) was added dropwise over approximately 15 minutes. The reaction continued to stir while warming to ambient temperature. After approximately 2 hr, the reaction solution was again cooled to 0-5 ^o^C and triethylamine (1.2 eq, 4.0 mL, 29 mmol) was added dropwise over approximately 10 minutes. To the stirring reaction mixture was added ice water (200 mL), solution transferred to separatory funnel, and product extracted into dichloromethane (500 mL). The dichloromethane layer was isolated and successively washed with ice water and brine, dried over sodium sulfate and concentrated to crude yellow colored oil. The product was purified by silica chromatography, eluting with 0-20% ethyl acetate/hexanes. Fractions containing product were combined, the solvent evaporated, and dried under high-vacuum at ambient temperature to afford **2-6** as a clear slight yellow colored oil. Yield: 6.13 g, 57%; ^1^H NMR (400 MHz, CDCl_3_) δ 8.08 (s, 1H), 7.40 – 7.27 (m, 15H), 4.89 (dd, *J* = 3.7, 1.4 Hz, 1H), 4.87 – 4.76 (m, 2H), 4.75 – 4.58 (m, 3H), 4.58 – 4.43 (m, 2H), 3.96 – 3.89 (m, 3H)

Perfluorophenyl 33-((2S,3S,4S,5S,6S)-3-acetamido-4,5-dihydroxy-6-(hydroxymethyl)tetrahydro-2H-pyran-2-yl)-18,18-bis(15-((2S,3S,4S,5S,6S)-3-acetamido-4,5-dihydroxy-6-(hydroxymethyl)tetrahydro-2H-pyran-2-yl)-5-oxo-2,9,12-trioxa-6-azapentadecyl)-16,23-dioxo-4,7,10,13,20,27,30-heptaoxa-17,24-diazatritriacontanoate (**2**) was synthesized analogously to compound **1** using (2S,3S,4S)-3,4-bis(benzyloxy)-2-((benzyloxy)methyl)-5-nitro-3,4-dihydro-2H-pyran (**2-6)** in place of (2R,3R,4R)-3,4-bis(benzyloxy)-2-((benzyloxy)methyl)-5-nitro-3,4-dihydro-2H-pyran in step 2 of the synthesis. LCMS m/z: 1776.9 [M+H]^+^; ^1^H NMR (400 MHz, D_2_O) δ 4.22 (dd, *J* = 10.9, 5.8 Hz, 3H), 4.15 – 4.07 (m, 3H), 3.99 – 3.93 (m, 5H), 3.90 (dd, *J* = 10.9, 3.3 Hz, 3H), 3.81 – 3.65 (m, 47H), 3.63 (t, *J* = 5.5 Hz, 6H), 3.58 (t, *J* = 6.4 Hz, 6H), 3.41 (t, *J* = 5.5 Hz, 6H), 3.09 (t, *J* = 5.8 Hz, 2H), 2.51 (t, *J* = 6.0 Hz, 8H), 2.03 (s, 9H), 1.88 – 1.66 (m, 6H), 1.63 – 1.42 (m, 6H); ^13^C NMR (101 MHz, DMSO) δ 170.31, 170.25, 169.37, 167.88, 73.33, 70.71, 70.24, 69.78, 69.74, 69.69, 69.67, 69.61, 69.57, 69.46, 69.34, 69.14, 68.24, 67.96, 67.42, 67.31, 66.84, 65.57, 59.87, 59.63, 50.27, 38.51, 36.58, 35.85, 33.96, 25.68, 22.62, 22.56; ^19^F NMR (376 MHz, D_2_O) δ -153.30 (d, *J* = 18.1 Hz, 2F), -158.09 (t, *J* = 21.8 Hz, 1F), -162.79 (dd, *J* = 22.0, 18.0 Hz, 2F); HRMS (*m/z*): [M+H]^+^ calculated for C76H127F5N7O34, 1776.8339; found, 1776.8322.

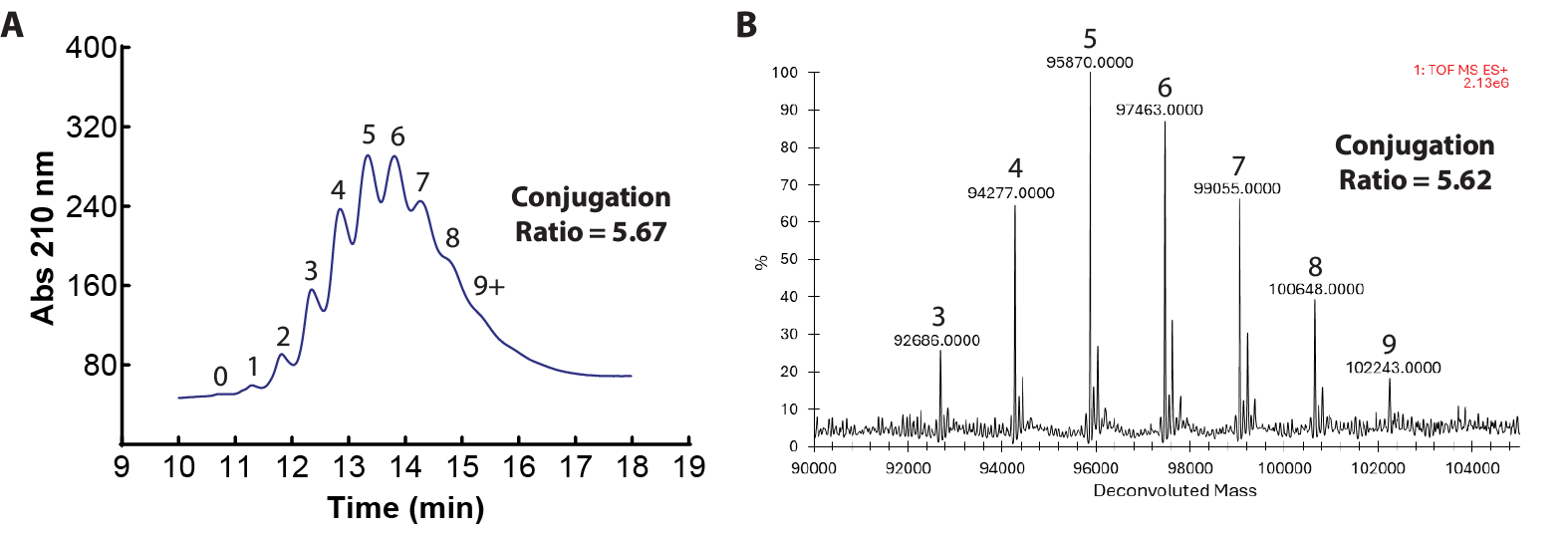

**Fig S1. MuSK LYTAC conjugation ratio analysis. (A)** Hydrophilic interaction chromatography and **(B)** mass spectrometry analysis of MuSK LYTAC conjugation ratio. Numbers above peaks denote the number of linker-ligands conjugated to MuSK Bait.

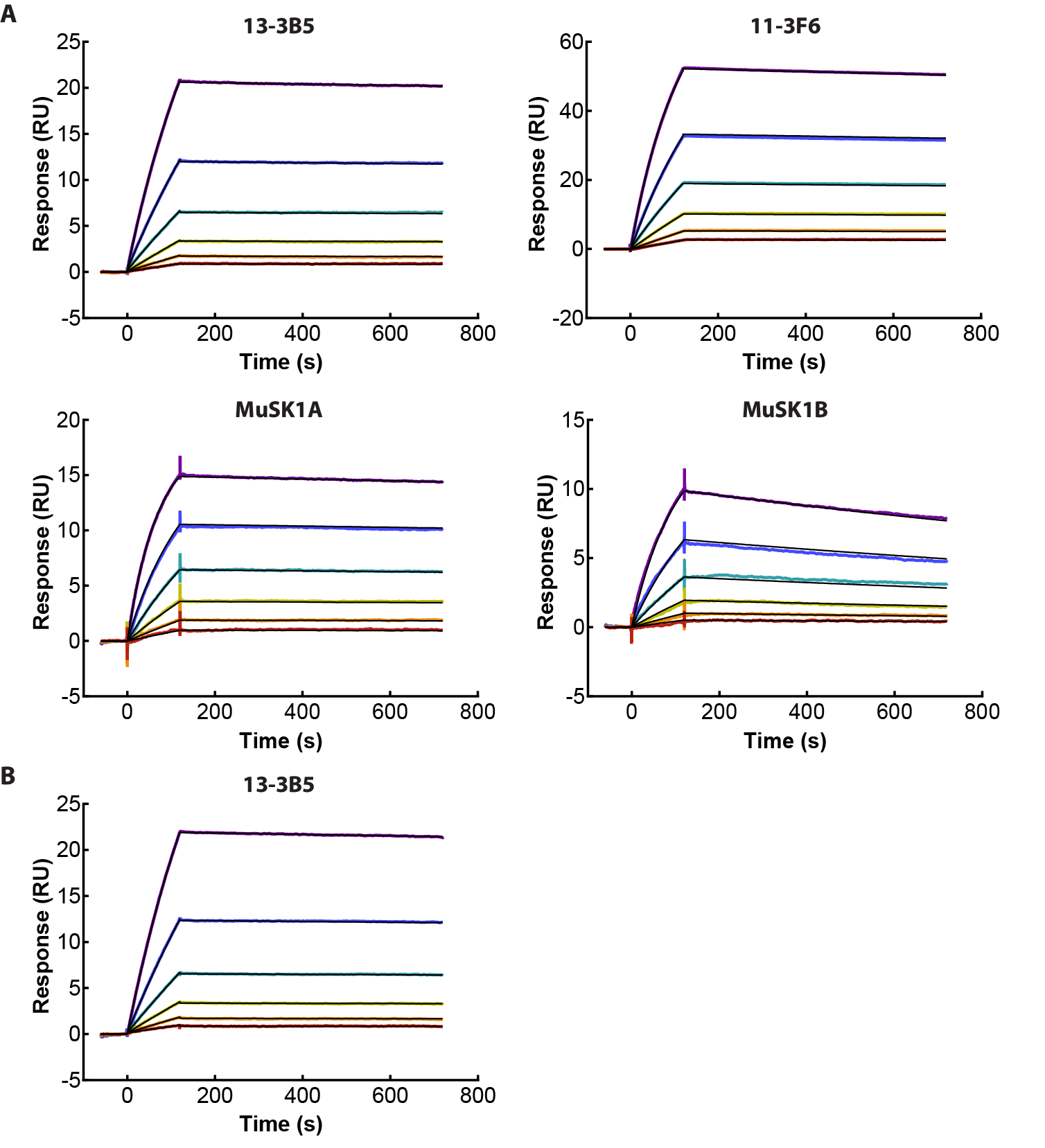

**Fig S2. MuSK Bait and MuSK LYTAC binding to immobilized MuSK antibodies.** Representative surface plasmon resonance (SPR) data with immobilized antibody, as indicated and varying concentrations of MuSK Bait **(A)** or MuSK LYTAC **(B)**. Average affinity values from three independent experiments are provided in Table 1.

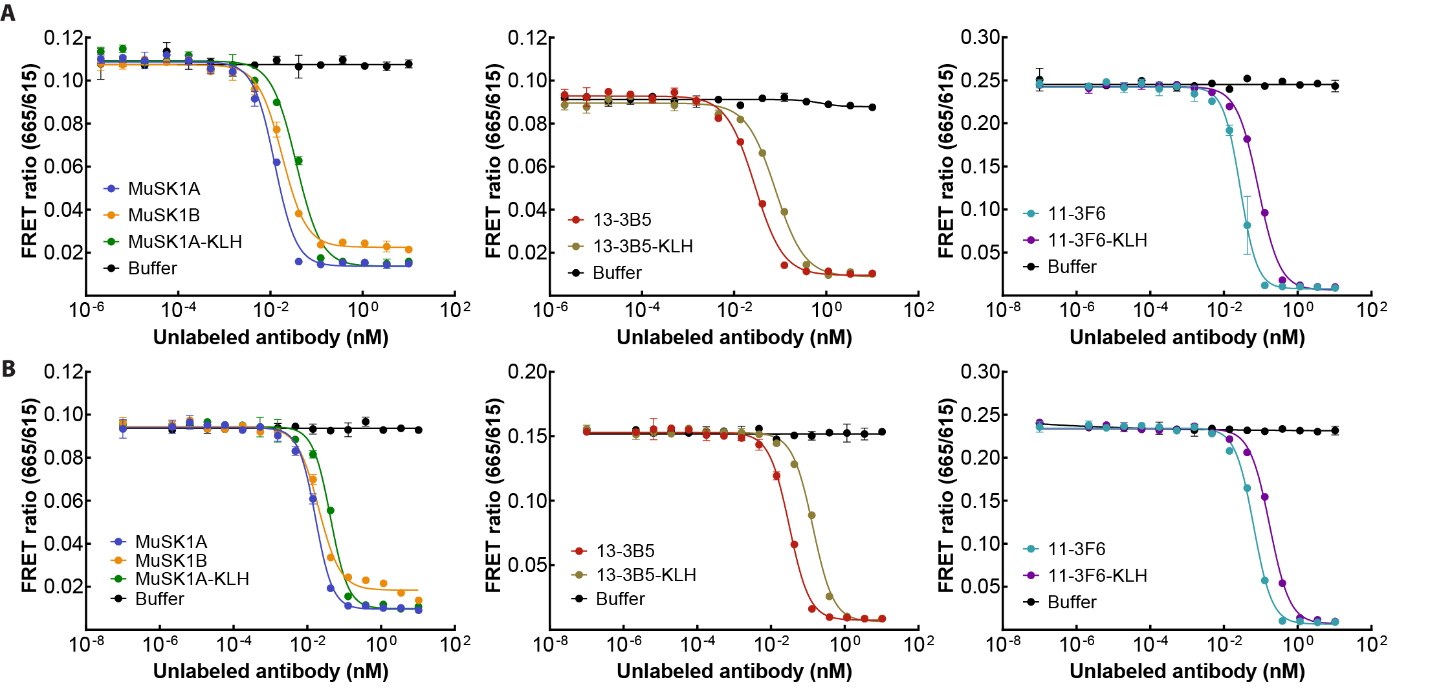

**Fig S3. MuSK Bait and MuSK LYTAC binding to MuSK antibodies in solution.** Representative time resolved fluorescence resonance energy transfer (TR-FRET) titrations curves with AF647-labeled MuSK1A and MuSK1B (left), 13-3B5 (middle) or 11-2F6 (right) with varying concentrations of indicated unlabeled antibody and MuSK Bait **(A)** or MuSK LYTAC **(B)**. Average affinity values from three independent experiments are provided in Table 1.

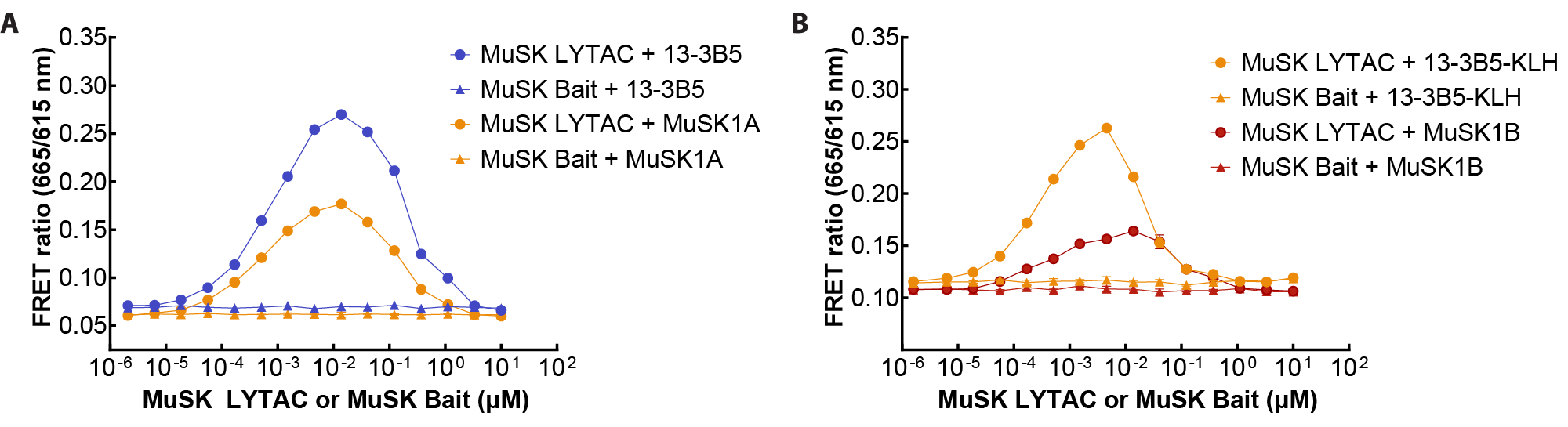

**Fig S4. MuSK LYTAC ternary complex formation.** TR-FRET-based ternary complex formation with recombinant ASGPR1, MuSK LYTAC or MuSK Bait, and AF647-labeled MuSK antibody **(A)** 13-3B5, MuSK1A, **(B)** 13-3B5-KLH, or MuSK1B. Data are representative of three independent experiments.

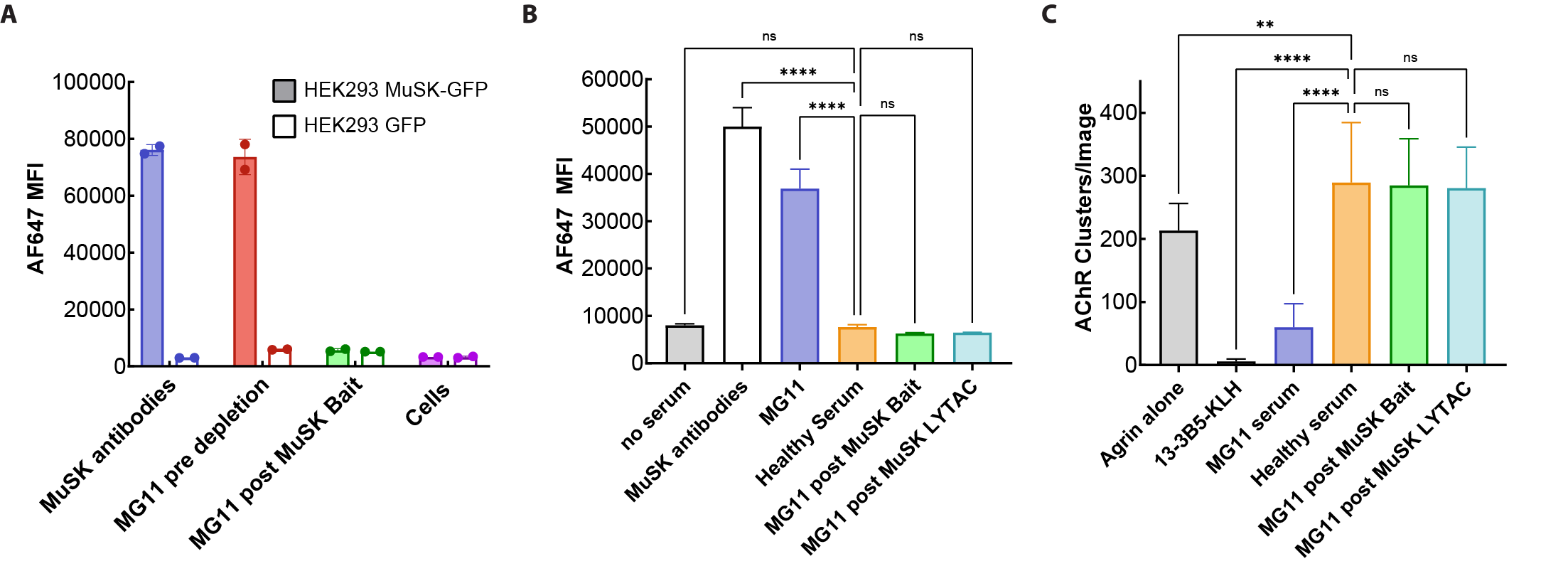

**Fig S5. MG patient serum depletion of MuSK antibodies. (A)** Cell binding assay with 3.0 µg/ml MuSK antibody mixture (1:1:1:1 11-3F6, 13-3B5, MuSK1A and MuSK1B) and MG11 patient sample pre and post depletion with MuSK Bait using HEK293 cells stably expressing MuSK-GFP or GFP. Cell binding assay **(B)** with MuSK-GFP-expressing HEK293 cells and AChR clustering assay **(C)** in C2C12 cells comparing effects of incubation of MG11 serum sample with MuSK Bait- or MuSK LYTAC-conjugated beads. Data are representative of at least three independent experiments.

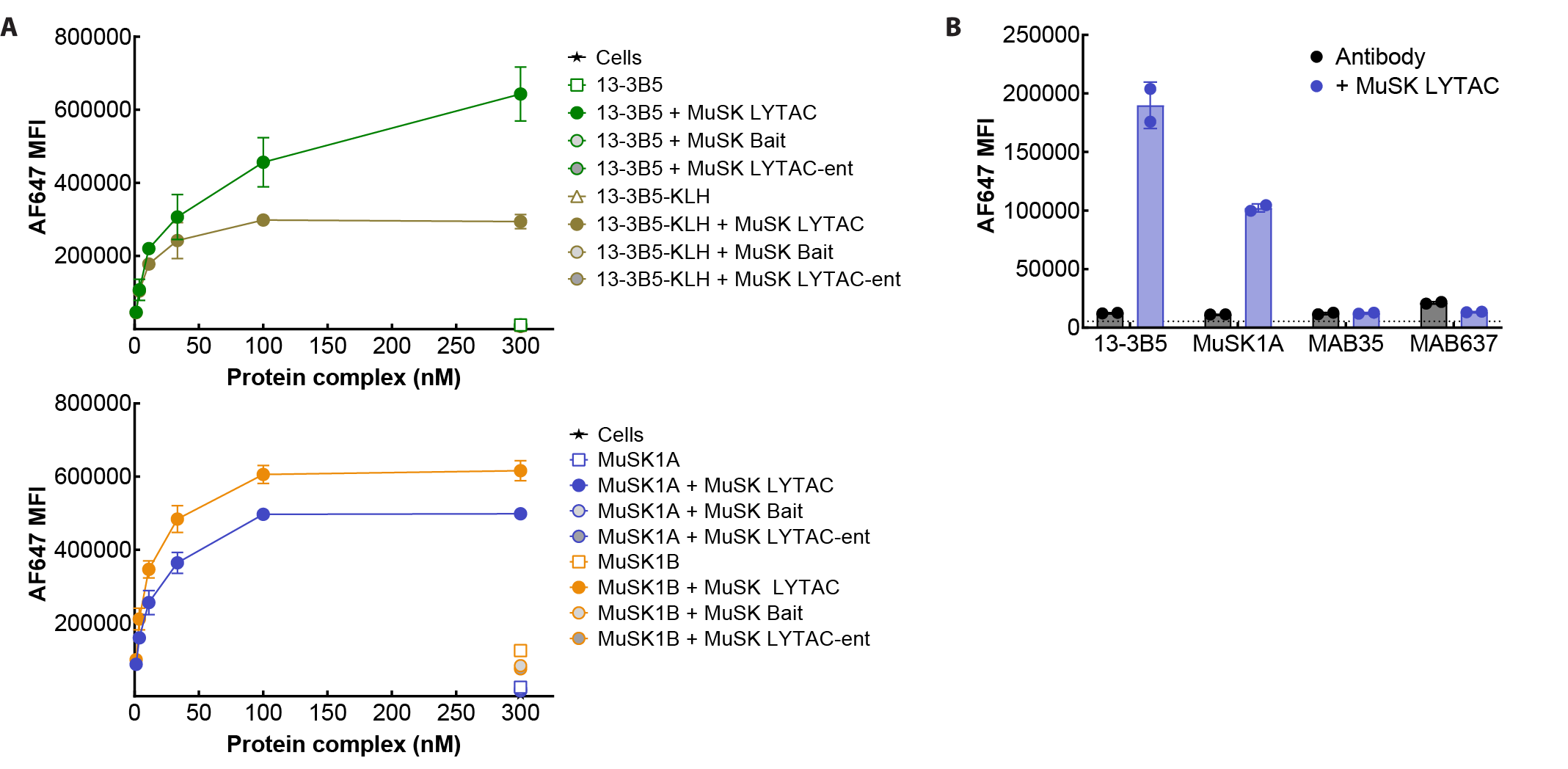

**Fig S6. LYTAC-dependent antibody cellular uptake.** **(A)** AF647-labeled MuSK antibody uptake with Hep G2 cells and MuSK LYTAC, MuSK Bait or MuSK LYTAC-ent, containing nonbinding, enantiomeric ASGPR ligands. **(B)** AF647-labeled MuSK antibody and AChR antibody uptake with MuSK LYTAC. MFI with cells alone denoted by dotted line.

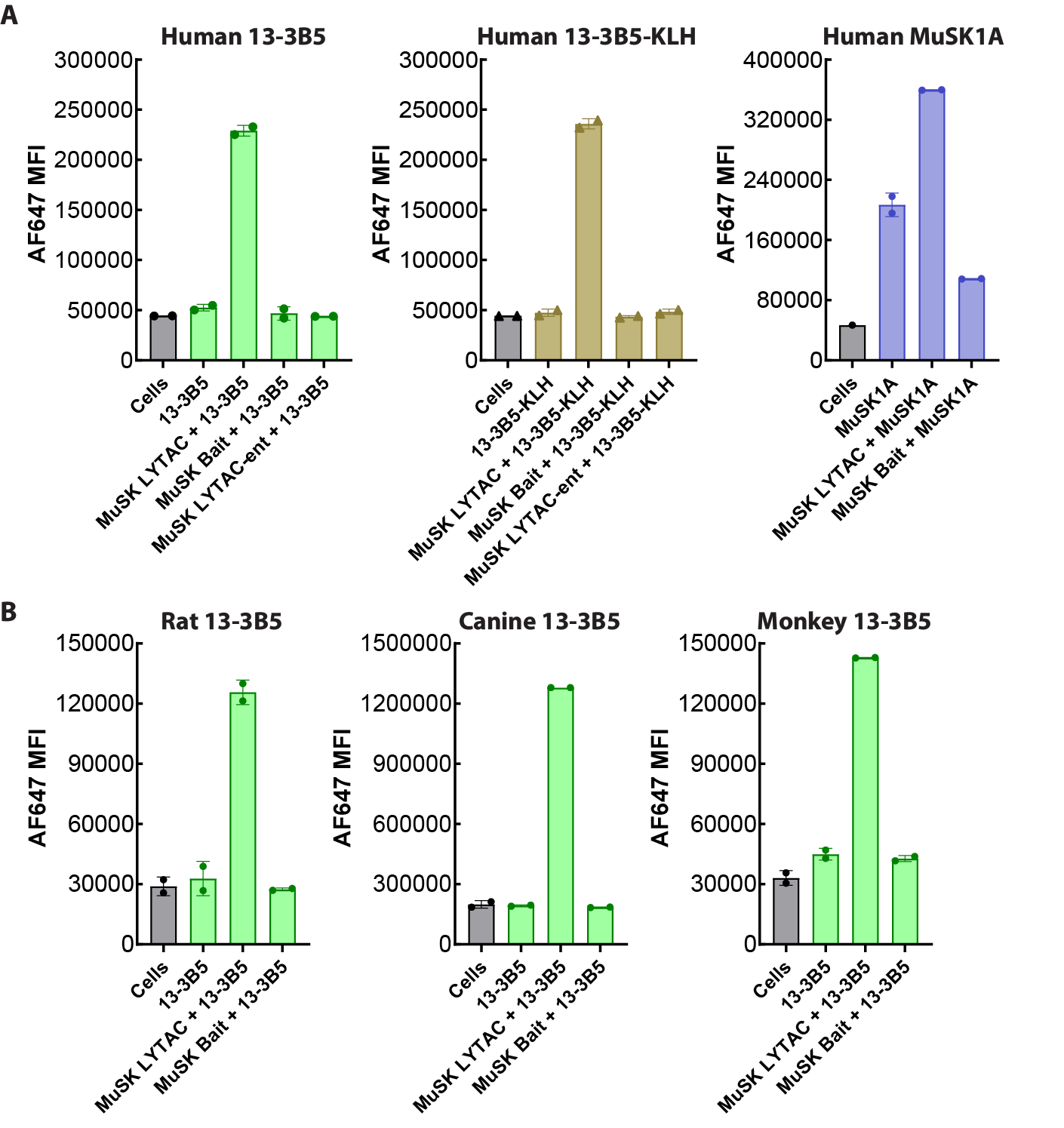

**Fig S7. MuSK LYTAC mediates antibody uptake in primary hepatocytes. (A**) AF647-labeled MuSK antibody uptake studies with 13-3B5 (left), 13-3B5-KLH (center) and MuSK1A (right) using primary human hepatocytes. Data is shown for cells alone, cells with the corresponding AF647-labeled antibody, or cells with antibody and MuSK LYTAC, MuSK Bait or MuSK LYTAC-ent. **(B)** AF647-labeled 13-3B5 cellular uptake with primary rat (left), canine (center) or cynomolgus monkey (right) hepatocytes with cells alone, cells with 13-3B5, or cells with antibody and MuSK LYTAC or MuSK Bait.

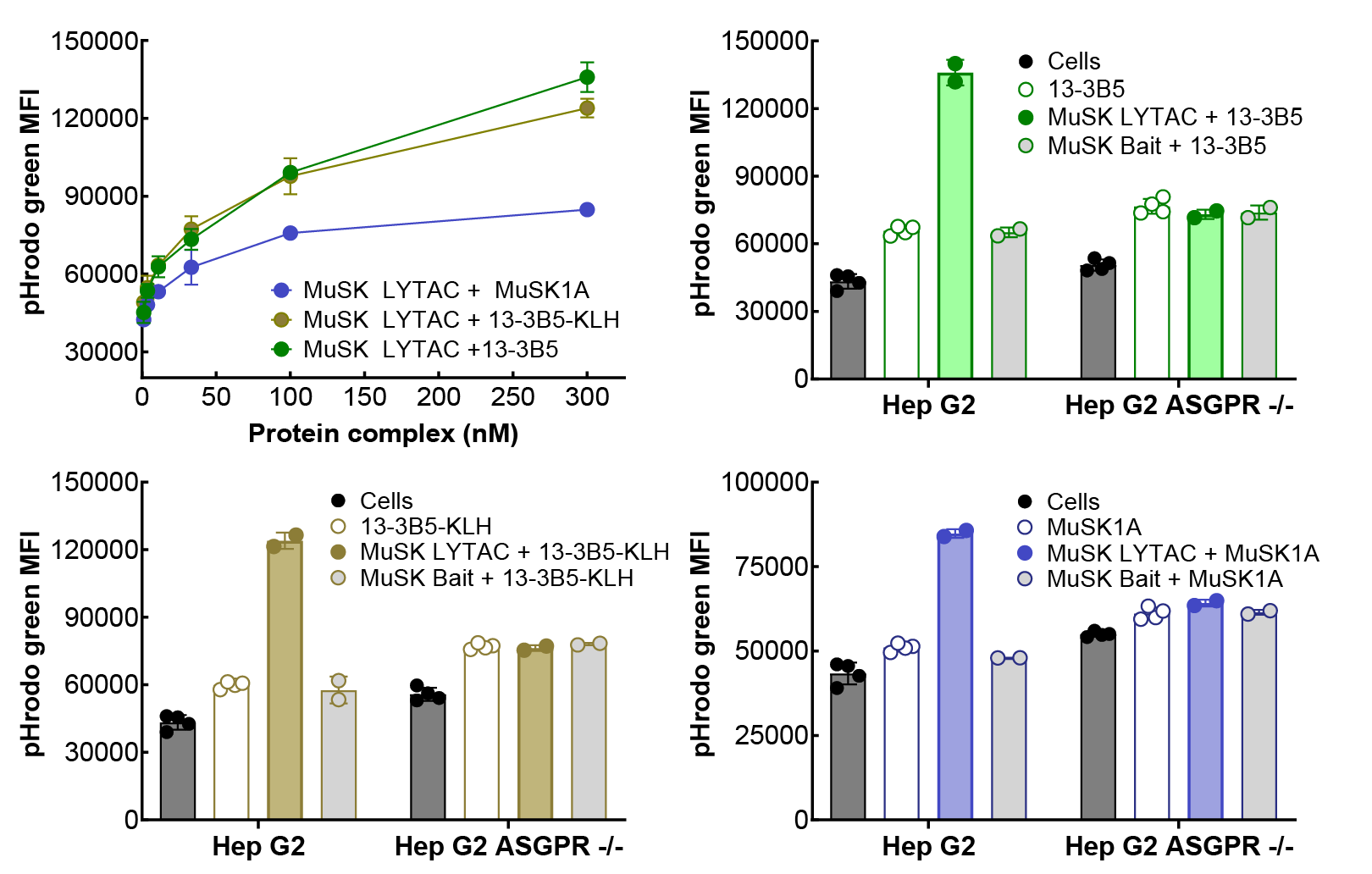

**Fig S8. LYTAC-mediated antibody delivery to acidic compartment in Hep G2 cells.** MuSK antibody acidic compartment accumulation measured by flow cytometry with pHrodo green labeled MuSK1A, 13-3B5 or 13-3B5-KLH shown as a function or protein concentration in Hep G2 cells (top left) and with cells alone or with 300 nM proteins in Hep G2 and ASGPR knockout (ASGPR -/-) Hep G2 cells (top right, bottom left and bottom right).

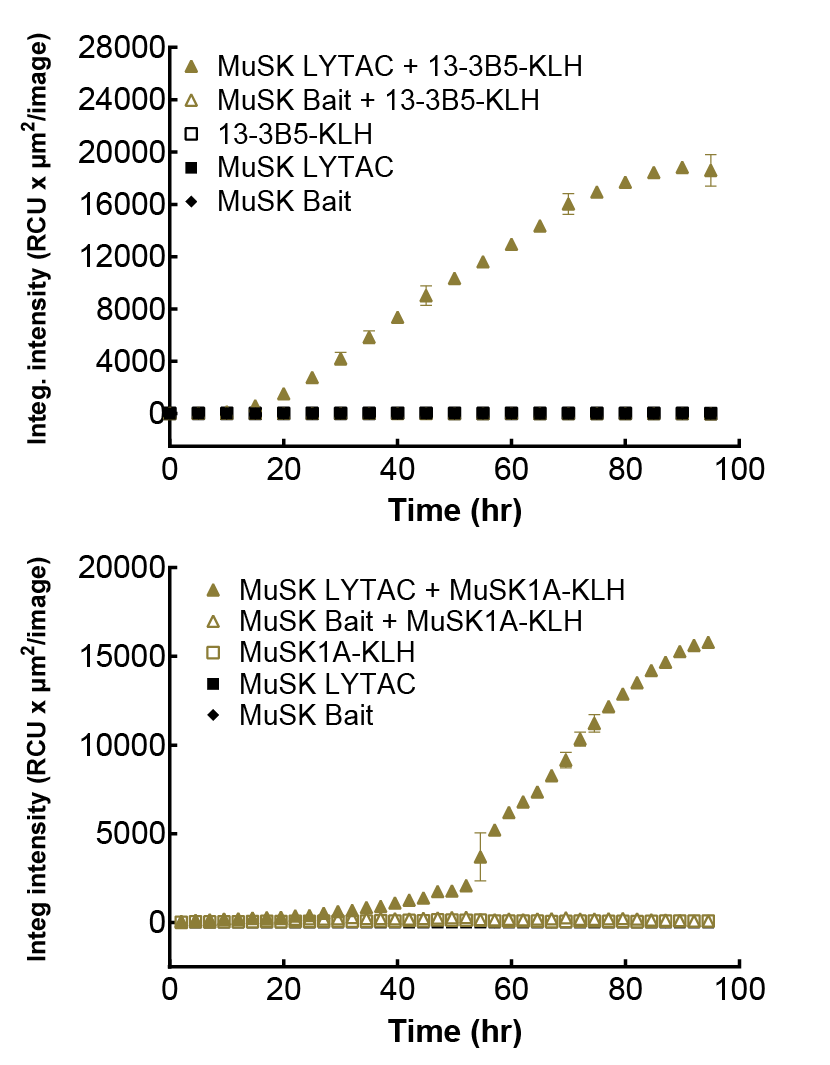

**Fig S9. LYTAC-mediated monovalent MuSK antibody degradation in Hep G2 cells.** 13-3B5-KLH (top) and MuSK1A-KLH (bottom) degradation using a TAMRA/QSY7-conjugated anti-human F(ab′)2. Intensity measured by microscopy is shown as a function of time. Probe mixed with MuSK Bait or MuSK LYTAC and the indicated MuSK antibodies at a 1:1:1 ratio (50 nM each).

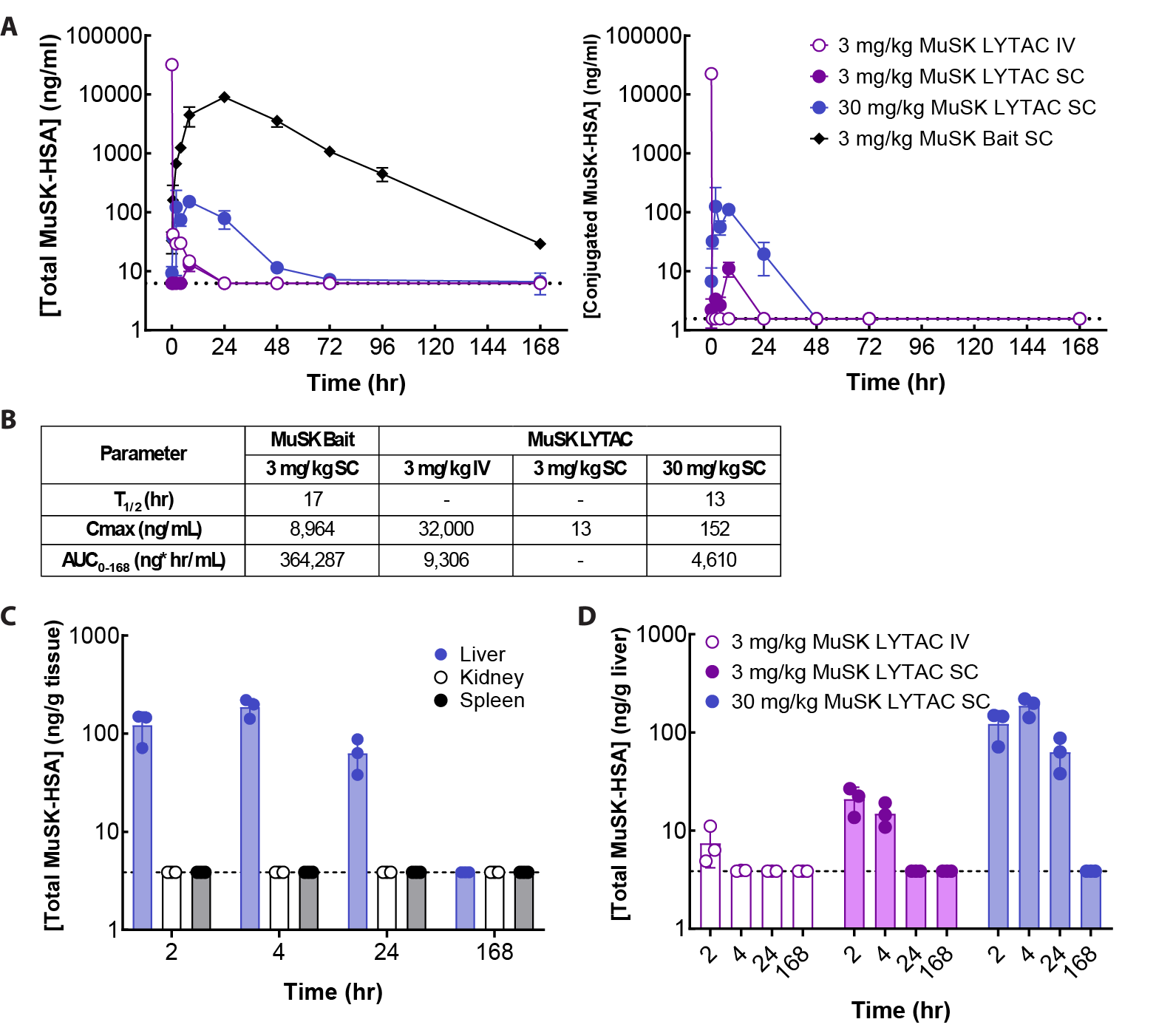

**Fig S10. MuSK LYTAC clears rapidly from serum and transiently accumulates in the liver of rat. (A)** PK profile of MuSK LYTAC administered IV or SC or MuSK Bait administered SC at the indicated dose levels in rat. Serum levels of total MuSK-HSA (left) or conjugated MuSK-HSA (right) were measured. The lower limits of detection were 6.25 ng/ml and 1.56 ng/ml for total and conjugate, respectively, as indicated by the dotted lines. **(B)** Serum PK parameters for total MuSK Bait or MuSK LYTAC. **(C)** Liver, kidney and spleen levels of MuSK LYTAC measured at the indicated timepoints after 30 mg/kg SC dosing. **(D)** Liver levels of MuSK LYTAC measured at the indicated timepoints after 3 mg/kg IV dosing or 3 mg/kg or 30 mg/kg SC dosing. Dotted line indicates ELISA limit of detection.

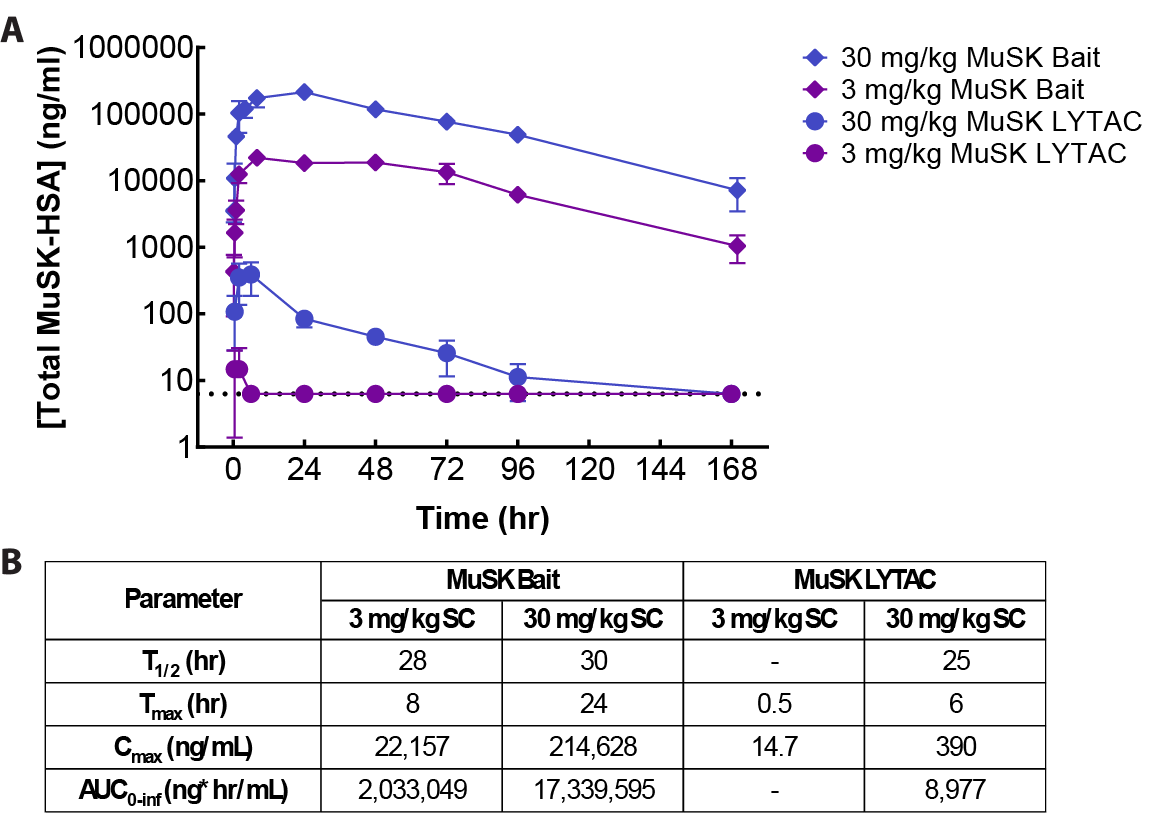

**Fig S11. Mouse PK of MuSK Bait and MuSK LYTAC.** Serum concentrations of total MuSK-HSA over time **(A)** and PK parameters **(B)** after SC administration of 3 mg/kg or 30 mg/kg of MuSK Bait or MuSK LYTAC. Dotted line indicates ELISA limit of detection.

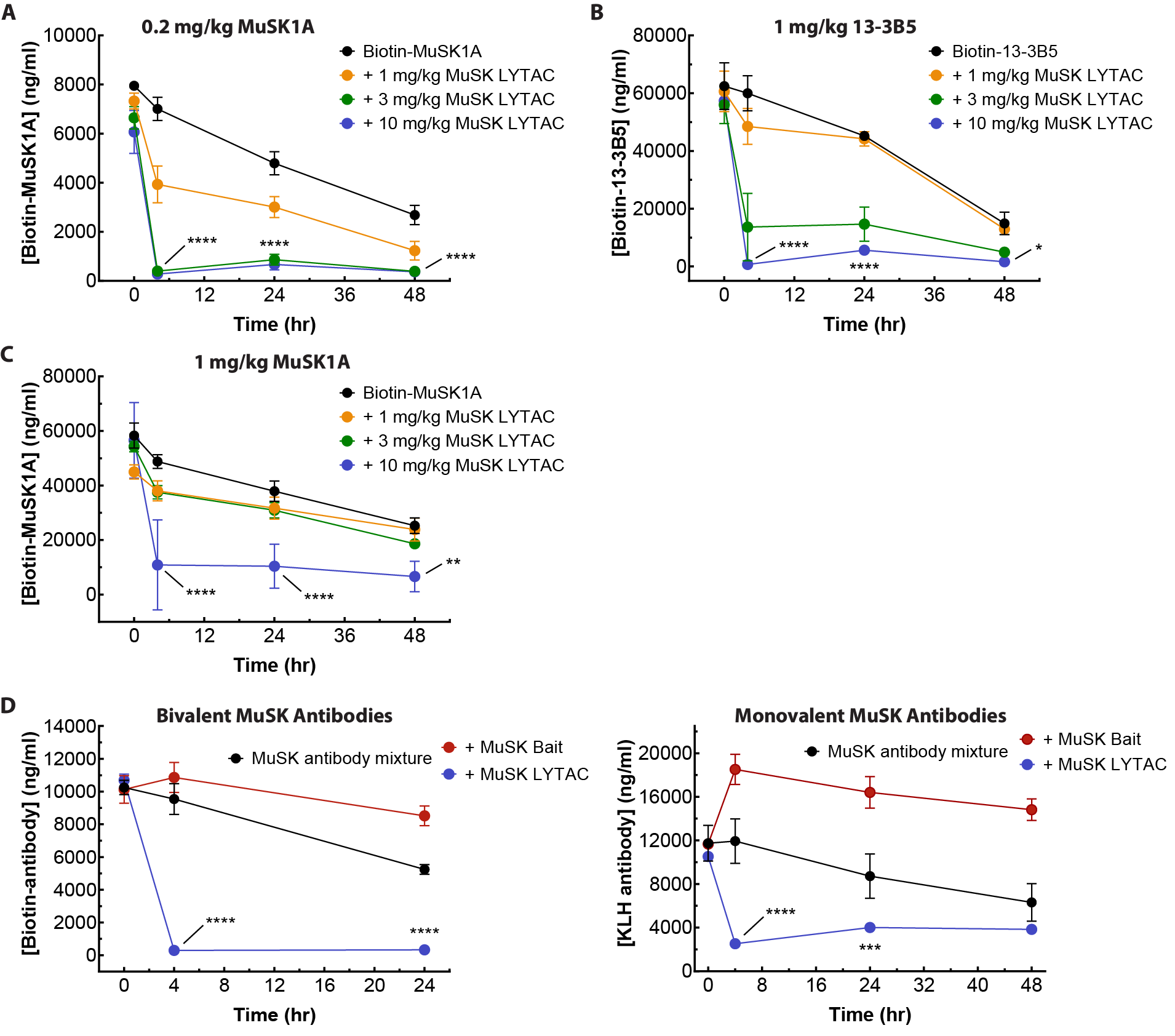

**Fig S12. MuSK LYTAC clearance of Ig1- and Ig2-binding MuSK antibodies.** Antibody levels (ng/ml) after 1, 3, or 10 mg/kg MuSK LYTAC SC treatment for **(A)** 0.2 mg/kg biotin-MuSK1A, **(B)** 1 mg/kg biotin-13-3B5, **(C)** 1 mg/kg biotin-MuSK1A. Bivalent or monovalent MuSK antibody levels (ng/ml) after 10 mg/kg MuSK LYTAC or 10 mg/kg MuSK Bait SC treatment for **(D)** 1 mg/kg total equimolar biotin-13-3B5 + biotin-11-3F6 + biotin-MuSK1A, or **(E)** 1 mg/kg kg total equimolar 13-3B5-KLH + 11-3F6-KLH + MuSK1A-KLH. Statistics: 2way ANOVA, comparison of 10 mg/kg MuSK LYTAC to Biotin-MUSK1A, Biotin-13-3B5 or MuSK antibody mixture, **** = P < 0.0001, *** = P < 0.0005, ** = P < 0.005, * = P < 0.05; n = 3 animals/group.

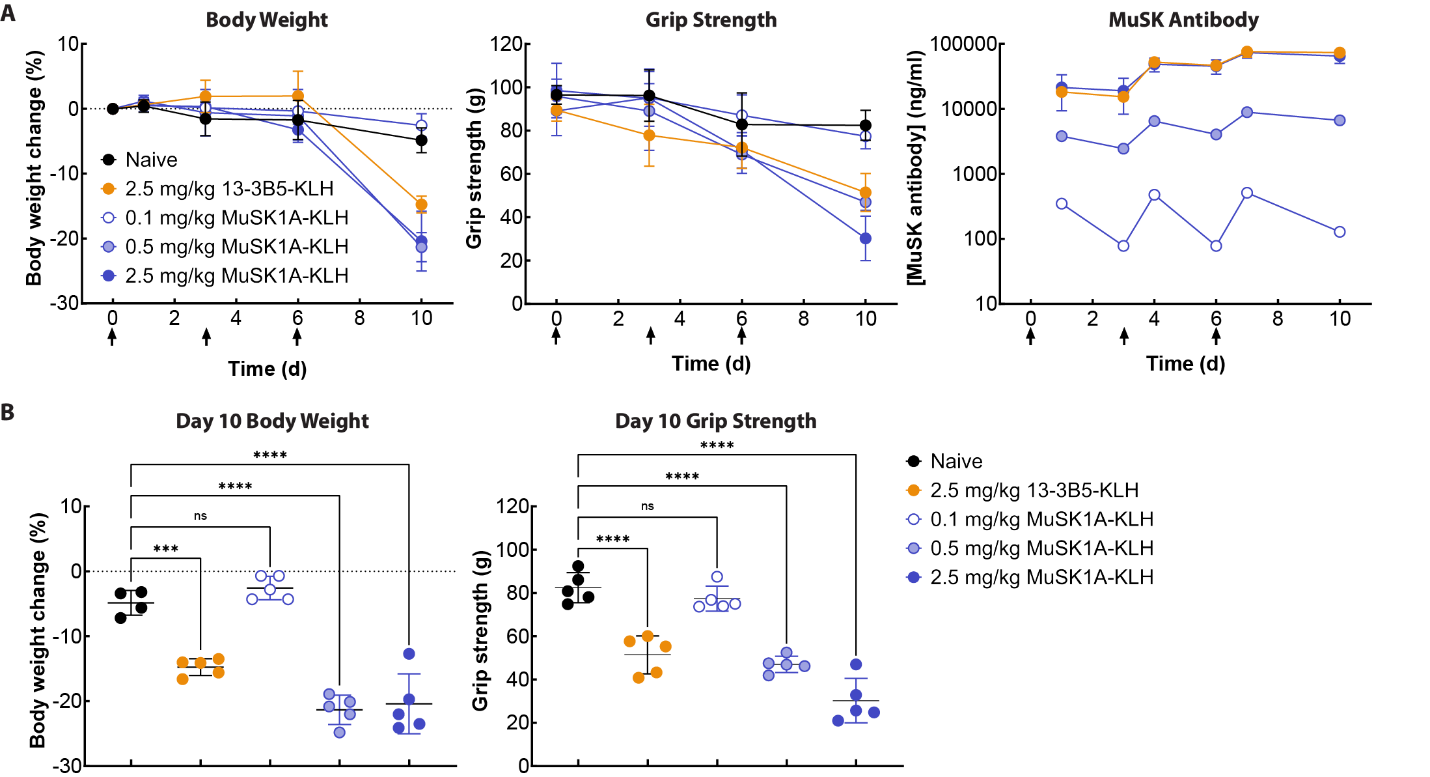

**Fig S13. Passive transfer of monovalent Ig2 domain binder MuSK1A induces weight loss and decreases grip strength in the mouse MuSK-MG model. (A)** Body weight (left), grip strength (middle), and antibody levels (right) measured after IP injection of the indicated amounts of MuSK1A-KLH on days 0, 3 and 6 (arrows). **(B)** Body weight (left) and grip strength (right) comparisons on day 10. One way ANOVA P value: **** <0.0001, *** <0.001; n = 5 animals/group.

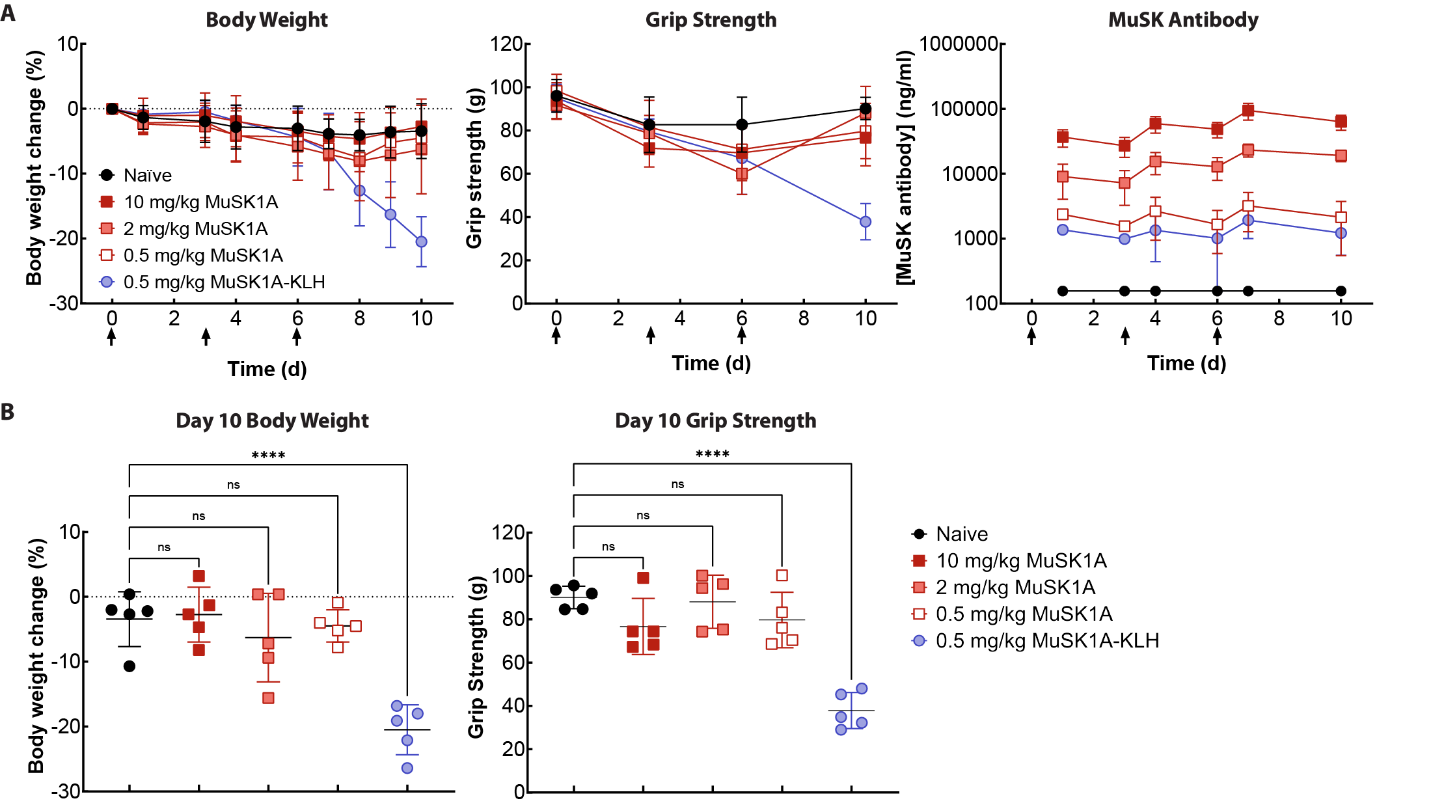

**Fig S14. Passive transfer of bivalent Ig2 domain binder MuSK1A does not induce disease symptoms in the mouse MuSK-MG model. (A)** Body weight (left), grip strength (middle), and antibody levels (right) measured after IP injection of the indicated amounts of MuSK1A or MuSK1A-KLH on days 0, 3 and 6 (arrows). **(B)** Body weight (left) and grip strength (right) comparisons on day 10. One way ANOVA P value: **** <0.0001; n = 5 animals/group.

**Table S1. Conjugation site mapping of MuSK LYTAC.**

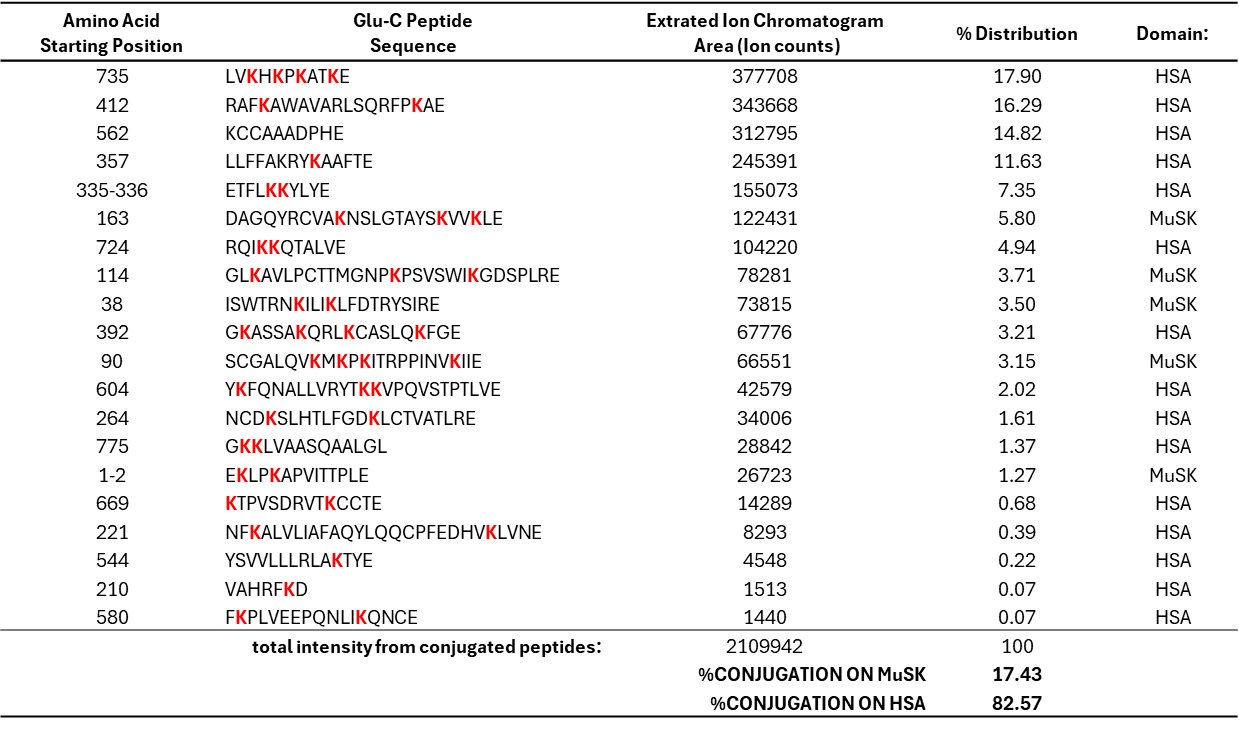

**Table S2. Patient clinical, laboratory and demographic data.**

| **Patient ID** | **Diagnosis** | **Serum MuSK Ab titer** | **Age** | **Sex** | **Disease duration**  **(years)** | **Treatment** | **MGFA class, MGC score at  TOC** | **Time since last RTX cycle**  **(months)** |
| --- | --- | --- | --- | --- | --- | --- | --- | --- |
| MG1 | Generalized MuSK MG | 8.05 | 25-29 | F | 3.1 | IVIG | IIa, 5 | - |
| MG2 | Generalized MuSK MG | 1.00 | 40-44 | F | 15 | None | 0, 0 | 2 |
| MG3 | Generalized MuSK MG | 8.02 | 30-34 | F | 12.9 | None | IIb, 4 | - |
| MG4 | Generalized MuSK MG | 0.73 | 20-24 | F | 3 | None | 0, 0 | 29 |
| MG5 | Generalized MuSK MG | 0.89 | 35-39 | F | 5 | MMF | IIb, 4 | - |
| MG6 | Generalized MuSK MG | 2.03 | 65-69 | F | 15.8 | None | 0, 0 | 2 |
| MG7 | Generalized MuSK MG | 0.82 | 45-49 | F | 12.5 | Pred | 0, 0 | 6 |
| MG8 | Generalized MuSK MG | 13.00 | 65-69 | M | 13.5 | Pred, MMF | 0, 0 | - |
| MG9 | Generalized MuSK MG | 7.69 | 45-49 | F | 6.2 | None | 0, 0 | 23 |
| MG10 | Generalized MuSK MG | 0.05 | 65-69 | M | 1.8 | Pred, AZA | IIa, 9 | 10 |
| MG11 | Generalized MuSK MG | + | 40-44 | F | 6 | Pred, MMF | Unknown | 5 |

Antibody titer was measured at the Mayo Clinic Laboratory (unit = nmol/L) for MG1-10. The cut off for negativity is ≤ 0. 02 nmol/L. TOC = time of collection; AZA = azathioprine; IVIG = intravenous immunoglobulin; Pred = prednisone; MMF = mycophenolate mofetil; RTX = rituximab; MGFA = Myasthenia Gravis Foundation of America; MGC = Myasthenia Gravis Composite.
